# Transmission of SARS-CoV-2 in standardised First Few X cases and household transmission investigations: a systematic review and meta-analysis

**DOI:** 10.1101/2022.04.01.22273107

**Authors:** Hannah C Lewis, Adrian J Marcato, Niamh Meagher, Marta Valenciano, Juan-Pablo Villanueva-Cabezas, Violeta Spirkoska, James E Fielding, Amalia Karahalios, Lorenzo Subissi, Anthony Nardone, Brianna Cheng, Soatiana Rajatonirina, Joseph Okeibunor, Eman A Aly, Amal Barakat, Pernille Jorgensen, Tasnim Azim, Pushpa Ranjan Wijesinghe, Linh-Vi Le, Angel Rodriguez, Andrea Vicari, Maria Van Kerkhove, Jodie McVernon, Richard Pebody, David J Price, Isabel Bergeri, the Unity Studies Collaborator Group

**Affiliations:** World Health Organization, Geneva, Switzerland; World Health Organization, Regional Office for Africa, Brazzaville, Congo; Department of Infectious Diseases, The University of Melbourne, at the Peter Doherty Institute for Infection and Immunity, Melbourne, Australia; Centre for Epidemiology & Biostatistics, Melbourne School of Population & Global Health, The University of Melbourne, Melbourne, Australia; Epiconcept, Paris, France; The Nossal Institute for Global Health, The University of Melbourne, Melbourne, Australia; Victorian Infectious Diseases Reference Laboratory, Royal Melbourne Hospital, at the Peter Doherty Institute for Infection and Immunity, Melbourne, Australia; School of Population and Global Health, McGill University, Montreal, Quebec, Canada; World Health Organization, Regional Office for the Eastern Mediterranean, Cairo, Egypt; World Health Organization Regional Office for Europe, Copenhagen, Denmark; World Health Organization, Regional Office for South-East Asia, New Delhi, India; World Health Organization, Regional Office for the Western Pacific, Manila, Philippines; World Health Organization, Regional Office for the Americas (Pan American Health Organization), Washington DC, United States of America; Murdoch Children’s Research Institute, Melbourne, Australia

**Keywords:** systematic review, meta-analysis, SARS-CoV-2, influenza, pandemic, transmission, household, epidemiological studies/investigation

## Abstract

We aimed to estimate the household secondary infection attack rate (hSAR) of SARS-CoV-2 in investigations aligned with the WHO Unity Studies Household Transmission Investigations (HHTI) protocol. We conducted a systematic review and meta-analysis according to PRISMA 2020 guidelines.

We searched Medline, Embase, Web of Science, Scopus and medRxiv/bioRxiv for ‘Unity-aligned’ First Few X cases (FFX) and HHTIs published between 1 December 2019 and 26 July 2021. Standardised early results were shared by WHO Unity Studies collaborators (to 1 October 2021). We used a bespoke tool to assess investigation methodological quality. Values for hSAR and 95% confidence intervals (CIs) were extracted or calculated from crude data. Heterogeneity was assessed by visually inspecting overlap of CIs on forest plots and quantified in meta-analyses.

Of 9988 records retrieved, 80 articles (64 from databases; 16 provided by Unity Studies collaborators) were retained in the systematic review and 62 were included in the primary meta-analysis. hSAR point estimates ranged from 2%–90% (95% prediction interval: 3%–71%; I^2^=99.7%); I^2^ values remained >99% in subgroup analyses, indicating high, unexplained heterogeneity and leading to a decision not to report pooled hSAR estimates.

FFX and HHTI remain critical epidemiological tools for early and ongoing characterisation of novel infectious pathogens. The large, unexplained variance in hSAR estimates emphasises the need to further support standardisation in planning, conduct and analysis, and for clear and comprehensive reporting of FFX and HHTIs in time and place, to guide evidence-based pandemic preparedness and response efforts for SARS-CoV-2, influenza and future novel respiratory viruses.

## Introduction

Coronavirus disease 2019 (COVID-19), caused by severe acute respiratory syndrome coronavirus 2 (SARS-CoV-2) infection, was first reported in Wuhan, China, in December 2019. It was declared a Public Health Emergency of International Concern by the World Health Organization (WHO) on 30 January 2020 and characterised as a pandemic in March 2020.^3^

Since the early stages of the COVID-19 pandemic, households have been a major setting of SARS-CoV-2 transmission.^4^ As such, household transmission investigations (HHTIs) – including studies of the First Few X cases (FFX) in household settings – provide an opportunity to explore transmission dynamics and infection-severity (i.e., probability of hospitalisation given infection) of SARS-CoV-2. HHTIs facilitate the collection of epidemiological, clinical, and virological data in well-defined closed settings, where household contacts can be more accurately ascertained and followed up to identify infector-infectee pairs than in the general population, including both virological and serological evidence of infection. Conducting HHTIs during the emergence of a novel respiratory pathogen provides an opportunity to swiftly characterise the transmissibility and infection-severity of the pathogen. These estimates are crucial to inform policy and public health interventions at the local and national level.

A systematic review and meta-analysis of within household transmission for 2009 pandemic influenza A(H1N1) found that estimates for household secondary infection rate/risk are highly heterogeneous.^5^ This was at least in part attributed to varying household definitions, secondary case ascertainment and testing methods, and duration of follow-up. This led to the recommendation of a unified approach for such investigations. Following a review of the global response to the 2009 A(H1N1) pandemic in 2012^6^, these recommendations were actualised through the global Consortium for the Standardization of Influenza Seroepidemiology (CONSISE)^7^ and the development of a suite of standardised early investigation protocols by WHO’s Global Influenza Programme and Influenza Pandemic Special Investigations and Studies (IPSS).^8^ These protocols were further adapted for other high threat respiratory pathogens, such as Middle East Respiratory Syndrome Coronavirus (MERS-CoV).^9^

With the emergence of SARS-CoV-2, the WHO quickly adapted the suite of standardised protocols for rapid use, re-branded them the Unity Studies^10^ and included protocols for FFX^11^ and HHTI.^12^ The Unity Studies protocols standardise methods to encourage rapid generation of local data for public health action and facilitate comparison of key epidemiological parameters, such as pathogen transmissibility and infection-severity, across regions and globally. They are adaptable to enable countries to conduct local investigations irrespective of income status and resource level.

We aimed to systematically review and meta-analyse available data from standardised FFX and HHTIs aligned with the objectives and methods of the WHO Unity Studies HHTI protocol for SARS-CoV-2 in order to: 1) describe the implementation of investigations in time and place 2) assess methodological quality of aligned investigations; 3) calculate a pooled estimate of SARS-CoV-2 household secondary infection and clinical attack rate and; 4) explore sources of heterogeneity in the household secondary infection and clinical attack rates. The data from these analyses will enhance overall understanding of the epidemiology of COVID-19 and inform future development of the HHTI protocol and implementation.

## Methods

The systematic review protocol was registered on PROSPERO on 5 August 2021 (registration number: CRD42021260065) and is reported according to the PRISMA 2020 guidelines.^13^

### Definitions

We defined Unity-aligned epidemiological SARS-CoV-2 HHTIs as investigations of index cases and all of their household contacts with longitudinal and prospective collection of epidemiological, virological and/or serological data for subsequent analysis.^12^ Retrospective investigations were considered aligned where the original data source pertained to contact tracing investigations with active follow-up of all household members of an index case.

Index cases were defined as the first case(s) of COVID-19 identified from a positive reverse transcription polymerase chain reaction (RT-PCR) result that subsequently triggered the recruitment of their household to the HHTI.

The WHO’s HHTI protocol defines a household as “[…] a group of people (two or more) living in the same residence […]”.^12^ In practice, the definition may vary across regions or when other aspects of livelihood are considered, such as income and the collective consumption of goods and services.^14^ We classified individuals who lived with index cases as ‘household contacts’. Only investigations with sufficient detail to characterise infection status of all household contacts were included in the systematic review and meta-analyses.

Our primary outcome, the household secondary infection attack rate (hereafter ‘hSAR’), was defined as the probability of a COVID-19 infection amongst susceptible household members exposed to the primary case detected using laboratory diagnostic tools (either RT-PCR or serology).^15^ The household secondary clinical attack rate (hereafter ‘hSCAR’) was our secondary outcome and defined as above using only clinical criteria. It was not always possible to distinguish between the index and primary cases within households, therefore we refer to primary or index cases as ‘index cases’.

### Search strategy

Three sources of data were used to identify records in our systematic review. First, four databases – MEDLINE, Embase, Web of Science and Scopus, were explored to identify relevant investigations published between 1 December 2019 and 26 July 2021. The search strategy sought to identify all combinations of “COVID-19” and terms relevant to HHTIs: “COVID-19” AND (“household transmission” OR “secondary attack rate” OR “close contact” OR “contact transmission” OR “contact attack rate” OR “family transmission”). The full search strategy is detailed in the Appendix.

Second, using the same search terms and dates, we screened unpublished investigations made available on the medRxiv and bioRxiv preprint servers. Finally, we screened results from FFX and HHTIs not yet available in the literature but that had been shared by Unity Studies Collaborators with WHO prior to 1 October 2021. We facilitated the registration of these investigations to the data repository “Zenodo”, with the permission of the Principal Investigators.^2^

### Eligibility criteria

Records were eligible for this review where the investigation:

1. was aligned to the WHO Unity Studies FFX and HHT investigation protocols, with longitudinally collected data and active follow-up of households with a RT-PCR confirmed index case.
2. reported at least five households recruited following identification of an index case.
3. reported the hSAR and/or hSCAR with a measure of uncertainty, or provided sufficient data to calculate these parameters.
4. was published in English, Chinese, French, Russian, Spanish, Portuguese, German or Italian.
5. represented an original investigation, and reported estimates from the most complete and relevant dataset available.

The search did not identify any records in languages other than those listed above.

### Screening and selection of articles

Records were imported into Covidence for consolidation, de-duplication and storage.^16^ Records were screened by title and abstract according to the eligibility criteria. Screening was performed by at least two independent reviewers (AJM, NM, JPV-C, VS, JEF) who were blind to the other reviewer’s assessment. A third independent reviewer (DJP) assessed records for eligibility where consensus could not be reached. Records in languages other than English, or where eligibility was unclear from initial screening, were retained for translation or full-text assessment.

Using the same methods for the abstract screening, we conducted full-text screening to determine the final eligibility of investigations. All records retained for the systematic review are henceforth referred to as investigations. Investigations that: 1) did not report sufficient information (e.g., full-text not available, conference abstract); 2) were secondary analyses of a previously included investigation or; 3) were investigations that included a significant proportion of vaccinated index cases or household contacts, were excluded.

### Methodological quality assessment

We used a flexible, fit-for-purpose critical appraisal tool that consisted of 12 items to assess the methodological quality of investigations.^17^ Briefly, the tool was developed to assess specific aspects of HHTI design, which combines features of case series and longitudinal studies. The tool builds upon well-established approaches to perform critical appraisal and risk of bias assessment of observational studies.^18,19^ Five team members (AJM, NM, JPV-C, VS, JEF) independently applied the methodological quality assessment checklist (two members per assessment) for the hSAR and hSCAR outcomes, and responses were recorded as Yes/No/Unclear. All questions were used for the overall assessment. In particular, questions related to household definition, secondary case ascertainment and duration of follow-up of households (Appendix) were highly relevant in our methodological quality assessment to understand alignment to the WHO Unity HHTI protocol and to indicate suitability in producing aggregated estimates of our outcome measures. Investigations were classified as having low, moderate, or high risk of bias for each outcome according to their methodological quality, i.e., investigations with lower methodological quality were more likely to have higher risk of bias. Where consensus about the methodological quality assessment could not be reached, a sixth reviewer (DJP/AK) finalised the assessment.

High risk of bias was generally attributed to unsuitable or ambiguous study design, analysis methods or results. This included, but was not limited to, unclear or broad definition of “household” and “household contacts”, unclear or unsuitable methods of secondary case ascertainment, unclear or insufficient follow-up duration, reduced exposure of household contacts to primary/index cases, or a combination of these.

### Data extraction

We extracted administrative and contextual data relevant to our study, including author, country, and timing of each investigation. We further contextualised each investigation by WHO region^20^, income status as reported by the World Bank in 2021^21^, and involvement in the United Nations Office for the Coordination of Humanitarian Affairs (OCHA) Humanitarian Response Plan (HRP) for COVID-19.^22^

We also collected total index cases, total households, total household contacts, total secondary cases, dates of investigation, household transmission design (i.e., was the primary objective of the study to characterise household transmission?), method of secondary case ascertainment (e.g., RT-PCR or serology, routine or symptom-based testing) and data collection methods (i.e., retrospective or prospective).

Data were extracted independently by two reviewers (AJM, NM, JPV-C, VS, JEF). Discrepancies in the data extraction were resolved by discussion with all participating reviewers. Where the reporting of estimates was unclear, incomplete, or not explicit, authors were emailed at least twice in November and December 2021 to confirm details. Responses were collated until 17 January 2022. Investigations were excluded if authors did not respond and sufficient data were not available. No restriction was put on the number of index cases per household. The number of households was assumed to be equal to the number of index cases if either was not reported or available upon follow-up, where the study design suggested this was a reasonable assumption.

### Statistical analysis

Values for SARS-CoV-2 hSAR, hSCAR and associated 95% confidence intervals (CIs) were extracted or calculated from crude data.

We conducted a meta-analysis to obtain a global estimate of SARS-CoV-2 hSAR. The primary meta-analysis only considered investigations assessed as having low or moderate risk of bias. Forest plots were produced to illustrate the hSAR and hSCAR estimates of included investigations. These are presented overall and by subgroups of interest as described below.

Heterogeneity was assessed by visually inspecting the overlap of the CIs on the forest plots, quantified in the meta-analysis using the I^2^ and 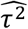 statistics, and assessed using the p-value from the χ^2^ test for heterogeneity.^23^ We fit a binomial-normal model to separately pool hSAR and hSCAR, as this model has been demonstrated to produce unbiased estimates and accounts for variation between investigations.^24^ I^2^ and 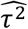 measures indicate the percentage of variation across investigations attributable to heterogeneity and the estimated between-investigation variance, respectively.

We further explored hSAR and hSCAR heterogeneity in pre-specified subgroup analyses. Subgroups included: 1) income setting (high income vs. low- and middle-income status) according to the World Bank classification^21^; 2) predominantly circulating variant at the time the investigation was conducted (variant of concern vs. other strains of SARS-CoV-2) according to data available from GISAID^25^ accessed via covariants.org^26^ where not reported in the investigations, and; 3) secondary case ascertainment methods (serological or RT-PCR testing of all household contacts vs. testing of symptomatic household contacts only). We also undertook a pre-specified subgroup analysis of hSAR using the results from our methodological quality assessment (low or moderate risk of bias vs. high risk of bias).

Following review of forest plots and meta-analysis results, 1) three post-hoc subgroups were defined and assessed in subgroup analysis and; 2) a post-hoc decision was made to report the 95% prediction interval for the primary analysis to further demonstrate the investigation-level heterogeneity. The first post-hoc subgroup analysis examined duration of follow-up of household contacts (14 days or less vs. greater than 14 days). The second compared investigations that did and did not comply with a stricter definition of adherence and alignment to the methods and objectives outlined in the Unity protocol. Strict methodological adherence was defined as a specifically designed household investigation with prospective follow-up and routine testing of all household contacts irrespective of symptoms. For example, investigations that only tested symptomatic household members would not meet this stricter methodological definition. The third subgroup compared investigations that used RT-PCR testing alone to those that used both RT-PCR and serological testing to ascertain secondary cases.

The results from initial and post-hoc analyses indicated substantial heterogeneity among included investigations which was not resolved by subgroup analysis. This led to an unplanned decision not to report any pooled estimated. As a result, several planned analyses, including assessment of publication bias and small study effect^27^, and a sensitivity analysis to investigate the effect of model specification^28^ were not undertaken (Appendix).

Data cleaning and collation was performed using Stata version 16 and R version 4.0.^29,30^. All pooled meta-analyses were undertaken in R version 4.0 using the *metafor* package.^31^

## Results

### Characteristics of included investigations

Figure 1 summarises the literature search and screening process. We identified 9,954 published records from database searches, and results from 34 FFX and HHTIs were provided directly from WHO Unity Studies Collaborators. Following removal of duplicates, 6,536 records went through title and abstract screening, and 284 subsequently underwent full-text assessment. A further 204 records were excluded at the full-text stage for various reasons (Figure 1). In total, 80 investigations (64 from database searches, and 16 provided by WHO Unity Studies Collaborators) were retained for data extraction, of which 62 (51 from database searches and 11 provided by WHO Unity Studies collaborators) were included in the primary meta-analyses.

**Figure 1.**
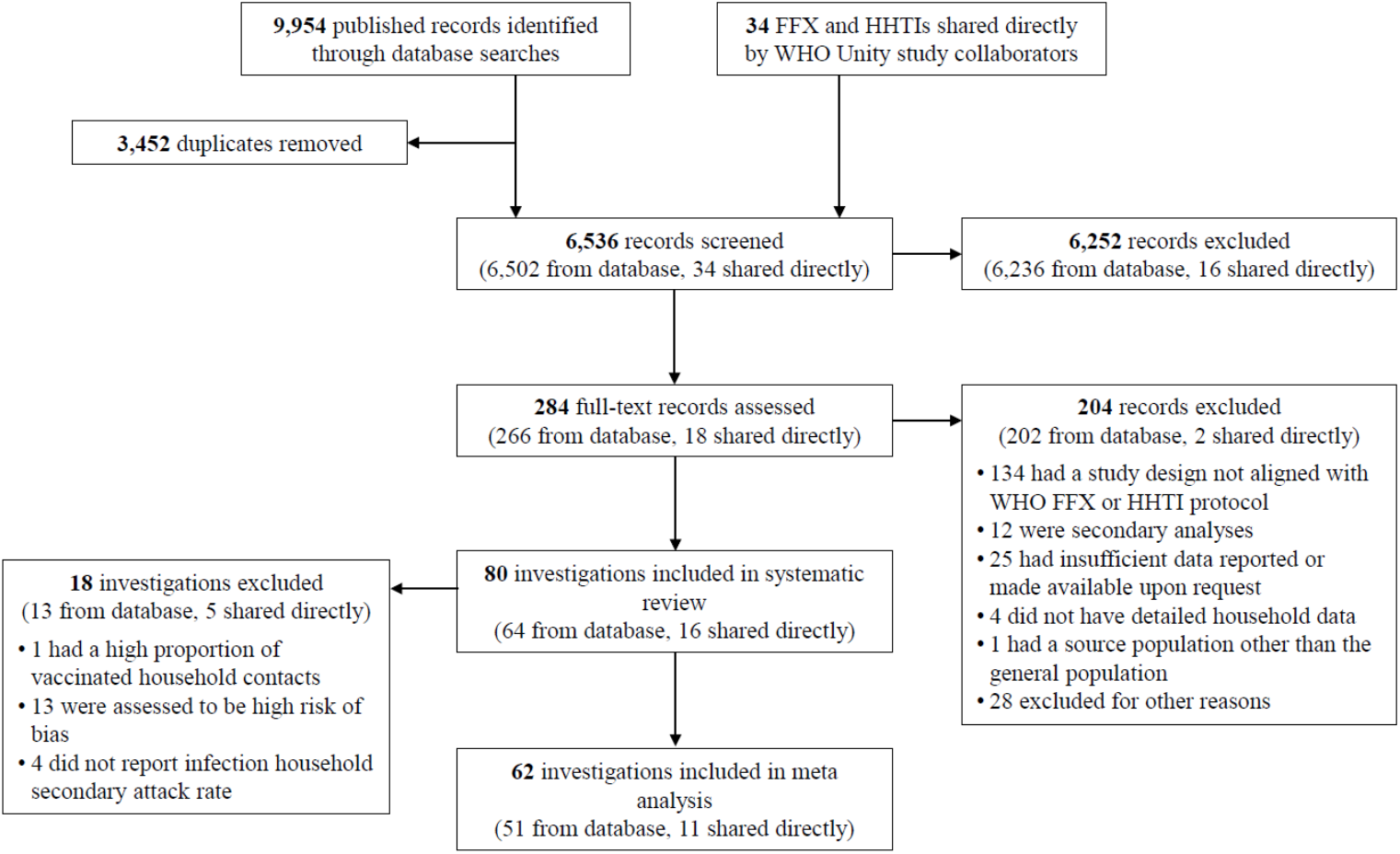
PRISMA flow chart. Other reasons for exclusion during full-text screening were: the full text was not accessible; duplicate investigations with different indexing; commentaries; corrections; non-COVID related investigations, and conference abstracts, preprints and short reports with subsequent publications. Abbreviations: FFX, First Few X cases investigations; HHTI, Household Transmission Investigation; WHO, World Health Organization.

Table 1 provides summary characteristics of the 80 investigations that met the eligibility criteria for the systematic review and for the subgroup of 62 articles that were included in the meta-analyses - detailed characteristics of each investigation can be found in Supplementary Table 1. Included investigations for the primary outcome of hSAR described follow-up of households between January 2020 and July 2021, with the majority (n = 50) completed before July 2020 (Supplementary Figure 1). Two investigations did not report a start date. Thirty-two countries were represented across all six WHO regions — fifteen countries contributed more than one investigation to this review, including 20 articles from China, 8 from India, 7 from the United States of America, 4 from South Korea, and 4 from Canada. Fifteen lower-and-middle income countries (LMICs) accounted for 51% (41/80) of the investigations identified in the systematic review, with 78% (32/41) of these being included in the meta-analyses. Nine investigations (11%) were conducted in countries supported in the OCHA HRP for COVID-19. Correspondingly, 17 high-income countries (HICs) accounted for 49% (39/80) of the investigations in the systematic review, with 77% (30/39) of these included in the meta-analyses.

**Table 1.** Summary statistics corresponding to all investigations included in the primary meta-analysis (n = 62) and in the systematic review (n = 80). Median and interquartile range [IQR, 25–75th percentiles] is reported for numeric quantities, and; number (%) is reported for categorical variables.

|  | Characteristic,<br>Median [IQR] or N (%) |  |
| --- | --- | --- |
|  | All investigations included in<br>meta-analyses<br>N = 62 | All investigations included in the<br>systematic review<br>N = 80 |
| Number of households included | 100.5 [44.5, 213.5] | 100 [38.75, 210.5] |
| Number of household contacts included | 286.5 [174.5, 768] | 279 [154, 792.75] |
| Total index cases | 103.5 [60.75, 259.75] | 102 [53.5, 229.5] |
| Secondary cases | 69.5 [37.75, 138.5] | 71 [34.75, 140.25] |
| **Source of article** |  |  |
| Peer-reviewed journal investigation –<br>identified in literature search | 48 (77.4%) | 58 (72.5%) |
| Preprints – identified in medRxiv and bioRxiv<br>literature search | 3 (4.8%) | 6 (7.5%) |
| Results provided directly from WHO Unity<br>Studies collaborators† | 11 (17.8%) | 16 (20.0%) |

| WHO Region‡ |  |  |
| --- | --- | --- |
| AFR | 3 (4.8%) | 6 (7.5%) |
| EMR | 1 (1.6%) | 2 (2.5%) |
| EUR | 13 (21.0%) | 18 (22.5%) |
| AMR | 9 (14.5%) | 13 (16.3%) |
| SEAR | 7 (11.3%) | 10 (12.5%) |
| WPR | 29 (46.8%) | 31 (38.8%) |
| Income Status§ |  |  |
| High | 30 (48.4%) | 39 (48.8%) |
| Upper-Middle | 20 (32.3%) | 23 (28.8%) |
| Lower-Middle | 9 (14.5%) | 13 (16.3%) |
| Low | 3 (4.8%) | 5 (6.3%) |
| Humanitarian Response Plan (HRP) for COVID-19 Status¶ |  |  |
| No | 58 (93.5%) | 71 (88.8%) |
| Yes | 4 (6.5%) | 9 (11.3%) |
| Presumed Dominant Strain†† |  |  |
| Variant of Concern | 4 (6.5%) | 6 (7.5%) |
| Other Strain | 58 (93.5%) | 74 (92.5%) |
| HHTI design |  |  |
| Yes | 35 (56.5%) | 43 (53.8%) |
| No (FFX, Case Series, Surveillance) | 27 (43.5%) | 37 (46.3%) |
| Method of Secondary Case Ascertainment (for hSAR)‡‡ |  |  |
| RT-PCR (irrespective of symptoms) | 34 (54.8%) | 43 (53.8%) |
| RT-PCR (symptom based) | 8 (12.9%) | 9 (11.3%) |
| RT-PCR or serology | 12 (19.4%) | 12 (15.0%) |
| Serology | - | 3 (3.8%) |
| Unknown method of lab diagnosis | 8 (12.9%) | 9 (11.3%) |
| Did not report a hSAR | - | 4 (5.0%) |

| Method of Secondary Case Ascertainment (for hSCAR)§§ |  |  |
| --- | --- | --- |
| Symptoms only | 24 (38.7%) | 33 (41.3%) |
| Did not report a hSCAR | 38 (61.3%) | 47 (58.8%) |
| Follow-up of households |  |  |
| Prospective | 44 (71.0%) | 55 (68.8%) |
| Retrospective | 17 (27.4%) | 23 (28.8%) |
| Unclear | 1 (1.6%) | 2 (2.5%) |
| Strict adherence and alignment to the Unity Study protocol¶¶ |  |  |
| No | 43 (69.4%) | 58 (72.5%) |
| Yes | 19 (30.6%) | 22 (27.5%) |
| Duration of follow-up |  |  |
| 14 days or less | 47 (75.8%) | 58 (72.5%) |
| Greater than 14 days | 11 (17.7%) | 14 (17.5%) |
| Unclear | 4 (6.5%) | 8 (10.0%) |
| Risk of bias in investigations reporting hSAR |  |  |
| Low | 24 (38.7%) | 25 (31.3%) |
| Moderate | 38 (61.3%) | 38 (47.5%) |
| High | - | 13 (16.3%) |
| N/A – hSAR not reported | - | 4 (5.0%) |
| Risk of bias in investigations reporting hSCAR |  |  |
| Low | 21 (33.9%) | 22 (27.5%) |
| Moderate | 3 (4.8%) | 11 (13.8%) |
| N/A – hSCAR not reported | 38 (61.3%) | 47 (58.8%) |
| † Include peer-reviewed literature and early results now available on Zenodo.<sup>2</sup><br>‡ WHO Region <sup>20</sup>: AFR (Africa), AMR (Americas), EMR (Eastern Mediterranean), EUR (Europe), SEAR (South East Asia), WPR (Western Pacific).<br>§ Income status as reported by the World Bank in 2021.<sup>21</sup><br>¶ Involvement in the United Nations Office for the Coordination of Humanitarian Affairs (OCHA) Humanitarian Response Plan (HRP) for COVID-19.<sup>22</sup><br>†† CoVariants (CoVariants: SARS-CoV-2 Mutations and Variants of Interest. (GISAID https://www.gisaid.org/<sup>25</sup> and https://covariants.org/<sup>26</sup>).<br>‡‡ hSAR (household secondary infection attack rate).<br>§§ hSCAR (household secondary clinical attack rate).<br>¶¶ Articles were defined based on strict adherence and alignment to the WHO Unity Studies HHTI protocol, including: (i) testing of all contacts (vs. only symptomatic), (ii) prospective data collection, (iii) specifically designed HH investigation (vs. FFX or surveillance data). |  |  |

Of the 76 investigations that reported a hSAR, secondary infections were predominantly ascertained by either scheduled RT-PCR testing of all contacts irrespective of symptoms (n = 43), symptom-initiated RT-PCR testing (n = 9), serology testing alone (n = 3), or through use of RT-PCR in combination with serology testing (n = 11). In the remaining 10 investigations, the method of secondary case ascertainment was unclear — these 10 investigations were excluded from the corresponding subgroup analysis. Secondary symptomatic cases were determined by symptoms alone in all instances where hSCAR was reported (n = 33).

Forty-three investigations (54%) were specifically designed as HHTIs. The remaining were investigations of all close contacts that reported sufficient detail from which we could estimate hSAR and/or hSCAR. The majority of articles collected data prospectively (n = 55), although some involved retrospective collation of detailed contact-tracing data from which households could be reconstructed (n = 23).

### Methodological quality assessment

The assessment of the methodological quality of the included investigations is summarised in Table 1 and detailed in the Appendix (Supplementary Figure 2 and 3). In total, 76 investigations reported a hSAR; 25 were considered to have a low risk of bias, 38 a moderate risk of bias, and 13 a high risk of bias. Of the 33 investigations reporting hSCARs, 22 were considered to have a low risk of bias and 11 had a moderate risk of bias.

### Household Secondary Attack Rate

Investigations that did not report a hSAR (n = 4), had a high risk of bias (n = 13) or were conducted in highly vaccinated cohorts (n = 1) were excluded from the main hSAR meta-analyses. In total, 18 investigations were excluded from the primary meta-analysis.

Point estimates for the hSAR (n = 62) ranged from 2% – 90% with a 95% prediction interval from 3% – 71% (Figure 2) and I^2^ = 99.7%, suggesting substantial heterogeneity between included investigations. The meta-analyses showed the heterogeneity between investigations was not reduced when examining the subgroups of interest (Table 2, Supplementary Table 2) — including by income setting; predominant circulating strain; testing protocol for household contacts, and; risk of bias. In all analyses, hSAR estimates varied substantially and I^2^ values were >99%. Due to this large amount of heterogeneity, pooled estimates of the hSAR are not reported. As a result of not producing any pooled estimates, evaluation of bias due to small study effect and sensitivity analysis to assess the effect of model choice were not carried out (Appendix).

**Figure 2:**
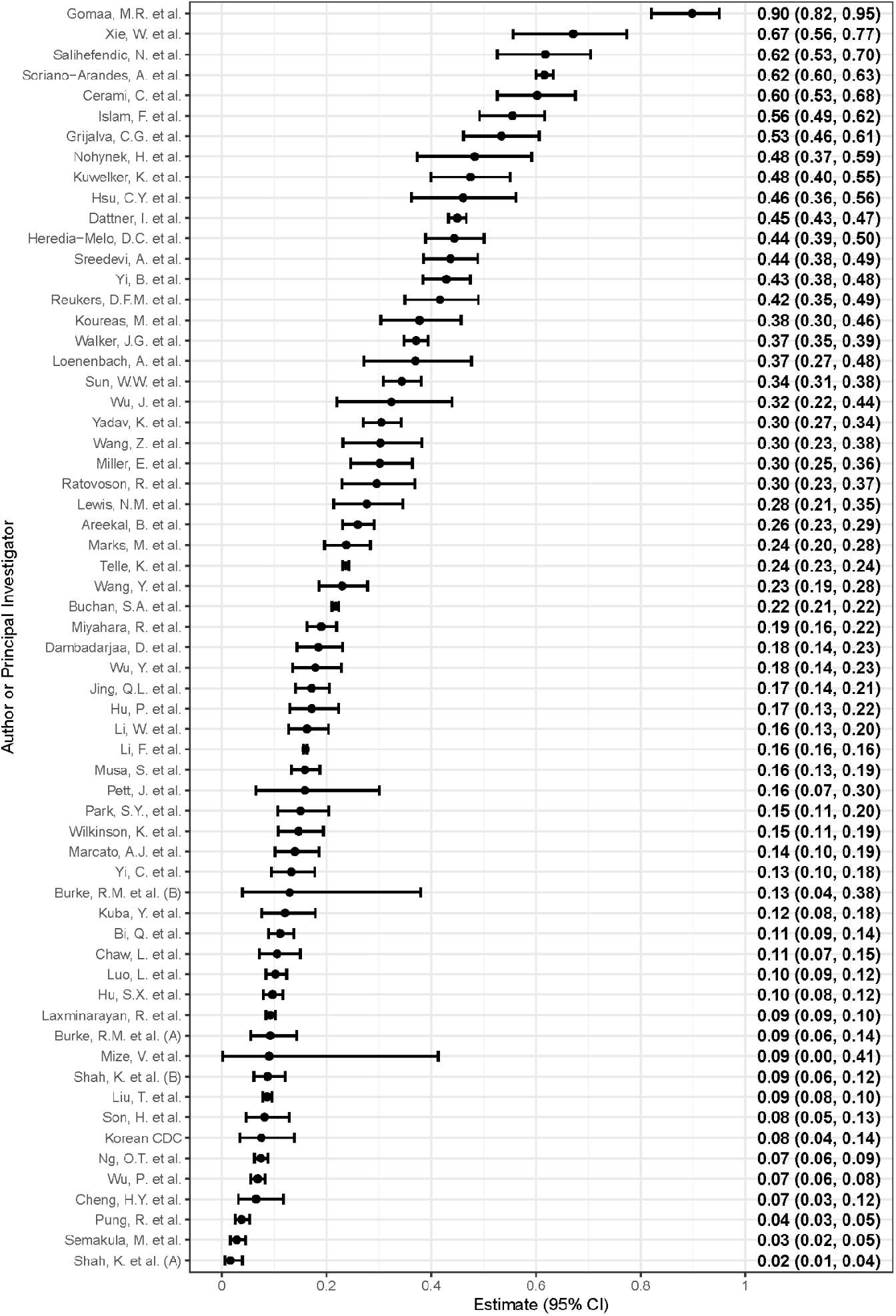
*Forest plot of the household secondary infection attack rates (hSAR) in included* investigations *(n = 62), ordered from highest estimated hSAR (top) to lowest estimated hSAR (bottom). The hSAR and 95% confidence intervals (CI) are shown on the right margin*

**Table 2.** Results from meta-analyses of household secondary infection attack rate (hSAR). I^2^ and 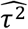 are presented for each model to indicate the percentage of variation across investigations attributable to heterogeneity and the estimated between-investigation variance, respectively. The p-value from the χ^2^ test for heterogeneity is also presented.

| | No. investigations | I<sup>2</sup> | $\widehat{\tau^2}$ | P-value |
| --- | --- | --- | --- | --- |
| Infection household secondary attack rate | 62 | 99.7 | 1.190 | <0.0001 |
| Pre-Specified Subgroup Analyses |  |  |  |  |
| Income Status† | 62 | 99.6 | 1.188 | <0.0001 |
| *High income* | 30 |  |  |  |
| *Low- and middle- income* | 32 |  |  |  |
| Predominant circulating strain‡ | 62 | 99.6 | 1.130 | <0.0001 |
| *Other strain* | 58 |  |  |  |
| *Variant of concern* | 4 |  |  |  |
| Testing protocol for household contacts | 54 | 99.4 | 1.174 | <0.0001 |
| *Testing of all contacts* | 46 |  |  |  |
| *Testing of symptomatic contacts* | 8 |  |  |  |
| Post-Hoc Subgroup Analyses |  |  |  |  |
| Strict adherence and alignment to the WHO Unity Studies protocol | 54 | 99.5 | 0.797 | <0.0001 |
| *Aligned* | 43 |  |  |  |
| *Not aligned* | 19 |  |  |  |
| Duration of follow-up of household contacts | 58 | 99.6 | 1.005 | <0.0001 |
| *14 days or less* | 47 |  |  |  |
| *Greater than 14 days* | 11 |  |  |  |
| Laboratory diagnosis method | 54 | 99.3 | 1.030 | <0.0001 |
| *RT-PCR only* | 42 |  |  |  |
| *RT-PCR and serology* | 12 |  |  |  |

† Income status as reported by the World Bank in 2021.<sup>21</sup>
‡ CoVariants (CoVariants: SARS-CoV-2 Mutations and Variants of Interest. (GISAID https://www.gisaid.org/ (24) and https://covariants.org/<sup>26)</sup>).

Supplementary Figures 5 – 11 show the forest plots by pre-specified and post-hoc subgroups (including adherence to the Unity protocol; duration of follow-up, and; use of serology to ascertain secondary cases), and Supplementary Table 2 examines the secondary meta-analysis to assess the effect of including high risk of bias investigations. All results indicate substantial heterogeneity. Supplementary Figure 4 shows a forest plot of hSAR estimates by WHO region, which was not examined in a subgroup meta-analysis due to correlation with income status.

Forest plots and meta-analysis of hSCAR are shown in Supplementary Figures 12 – 14 and Supplementary Table 3. As with the hSAR data, a high amount of heterogeneity between investigations was evident, which was not explained by any of the subgroups investigated. Consequently, pooled estimates of hSCAR were not produced.

## Discussion

This is the first systematic review and meta-analysis of standardised FFX and HHTIs – aligned with the objectives and methods of the WHO Unity Studies HHTI protocol – for SARS-CoV-2. We identified 80 investigations from 32 countries, 51% (41/80) of which were from 15 LMICs, showing that implementation was feasible in a range of settings and in every WHO region. Although all investigations were classified as aligned with the WHO Unity Studies protocol based on the information provided, we found substantial heterogeneity in the reported secondary attack rates (hSAR range: 2% – 90%; 95% prediction interval [PI]: 3% – 71%; I^2^ = 99.7). As a result, reporting of pooled estimates of hSAR or hSCAR was not deemed appropriate. Further subgroup analyses were undertaken to understand this heterogeneity, yet a similar extent of heterogeneity (I^2^ values were >99%) was still observed — consequently, pooled estimates are not provided.

The high degree of heterogeneity and wide range of estimates in reported hSAR is consistent with those of other recent systematic reviews and meta-analyses: one study reported an I^2^ value of 99.4% with hSARs ranging from 0% – 74%^32^ and another reported an I^2^ of 97.4%, with hSARs ranging from 4% – 55%.^33^ Nevertheless, these reviews reported pooled hSAR estimates of 18.9% (95% CI: 16.2% – 22.0%)^32^, 21.1% (95% CI: 17.4% – 24.8%)^28^ and 18.1% (95% CI: 15.7% – 20.6%).^33^

In the context of quantifying transmission, relevant potential sources of heterogeneity include methodological differences such as study design and definition of household, as well as differences in study context, such as current public health and social measures (PHSM), and population or individual behaviours. The I^2^ value estimates the proportion of the variance in reported estimates that is due to heterogeneity^34^, and should not be interpreted in isolation. When conducting meta-analyses, sources of heterogeneity should be identified *a priori* and explored in subgroup analyses where sufficient data have been reported.^35^ For example, Thompson^28^ identified a substantial difference in household attack rates when considering the reported duration of exposure between contact and case. This emphasises the need to precisely report the study design, epidemic context, and household dynamics, to meaningfully quantify transmission. Where available, we extracted information on these potential sources of heterogeneity, however sufficient detail on all aspects of the study design and implementation were not routinely or consistently reported.

Visual inspection of the forest plots suggests potentially lower hSAR in the Western Pacific Region (WPR) compared to other regions (Supplementary Figure 4). Most investigations conducted in the WPR were conducted during the first half of 2020. Several countries in the WPR enacted stringent PHSM and behavioural responses in early 2020, which resulted in low SARS-CoV-2 circulation.^32,36^ Furthermore, household size and structure (e.g., high-rise apartments, high- vs low-density households) varies between regions and urban-rural localities, as well as socio-economic status of different subpopulations which may all contribute to lower observed hSAR.^37^

The timing of each investigation must be considered in relation to local epidemic activity and evolving PHSM when interpreting the results reported in this meta-analysis (Supplementary Figure 1), however, these details were not sufficiently reported in included investigations. Most included investigations (n = 48) were finalised in the first six months of 2020, likely during circulation of the ancestral virus and early SARS-CoV-2 variants, i.e., prior to the designation of Alpha and Delta as Variants of Concern^38^ (Appendix, Supplementary Figure 1), which are known to have increased transmissibility.^39,40^ Only six investigations in the systematic review were conducted during a period when Alpha and/or Delta variants of concern were the dominant (or equally dominant) circulating strains.^25,26^ Among the six studies conducted after the designation of Alpha and Delta, two were excluded from the meta-analysis and three were conducted in the same country — providing insufficient representation for a subgroup analysis.

The frequency and type of specimen collection, as well as the duration of follow-up and laboratory methods employed, can influence the hSAR estimate. In our review, we observed high variability in the laboratory methods used to ascertain secondary cases, including the use of RT-PCR, serology or both. However, we found no difference in observed variability in post-hoc subgroup analyses. We also explored heterogeneity in hSAR estimates by duration of follow-up and whether all household contacts were tested to determine the influence of these study design aspects. Both of these analyses showed high variability across investigations. In addition, some investigations did not exclude non-susceptible individuals who tested positive by serology at baseline which may underestimate hSAR, although this information was inconsistently reported across investigations.

To assess the methodological quality of HHTIs, previous reviews adapted existing appraisal tools^18,19^ — however, these tools are limited as they were not designed for this purpose. We conducted a robust and thorough assessment of the methodological quality of investigations using a bespoke quality assessment tool for HHTIs which allowed for a more targeted critical appraisal and better understanding of strengths and limitations of HHTIs.^17^ We only included investigations with a low or moderate risk of bias in our primary meta-analysis (n = 62) and subsequently only those aligned with the objectives of the WHO Unity Studies HHTI protocol. It also allowed us to carefully consider the appropriateness of pooling data across investigations that were conducted in different resource settings and environments, using different protocols and with differing internal and external validity.

The use of a bespoke tool for methodological quality assessment does not guarantee that the intricacies of HHTI designs are fully captured, particularly where insufficient details are reported. We acknowledge that tailoring of the HHTI protocol according to cultural norms (e.g., household definition), capacity (e.g., laboratory testing, degree of follow-up) and context (e.g., PHSM, local incidence, quarantine practices) may be required, increasing the true variance observed across investigations. As a result, unclear reporting may have inflated the heterogeneity in our review due to inappropriate inclusion or exclusion of some investigations.

Of the 76 investigations that reported a hSAR, 13 were assessed to be at high risk of bias and subsequently excluded from the primary analyses. The assessment of methodological quality was strongly influenced by a range of factors, including: unclear or broad definition of household or household contacts (e.g., those that included more than residential contacts); unclear, unsuitable or incomplete laboratory and follow-up methods in HHTIs (e.g., symptom-based testing), and management of index cases that reduced exposure of household contacts to index cases (e.g., isolating index cases either within or outside the household).

Although the above methodological and contextual factors were deemed plausible sources of heterogeneity, none could explain a substantial amount of the variance in reported hSARs. This demonstrates the necessity for high-quality, standardised investigations and clear and comprehensive reporting of study design and household dynamics, to meaningfully quantify transmission. Further, in the presence of substantial contextual differences, we question the suitability of providing single pooled estimates of such pathogen characteristics. The first step in any meta-analysis is to consider whether the studies are all estimating the same quantity, and whether they should be pooled. Here, we have identified various sources of heterogeneity across settings which suggests they should not be pooled. While current reporting practices do not allow us to interrogate these heterogeneities further, future investigations based on consistent reporting guidelines may allow for more nuanced analyses where results can be pooled at an appropriate scale. In contrast, within a given country setting, many of the identified sources of heterogeneity are likely to be consistent across investigations (e.g., household structure), or, of sufficient relevance to the interpretation that they should be reported alongside investigations repeated over time (e.g., prior infection or vaccination histories). In these cases, sequential hSAR estimates from the same population over time, can provide crucial insight into vaccine program impacts and key characteristics of a novel pathogen (e.g., changes in transmissibility of SARS-CoV-2 variants) to help guide both national^41^ and international^42,43^ pandemic response and preparedness efforts.

While HHTIs from LMICs are underrepresented in the literature, this review included results from 41 investigations, of which 11 (27%) were shared by WHO Unity Studies collaborators at the end of February 2022 prior to peer review or pre-print publication. Such data would not typically be available for inclusion in a systematic review and meta-analysis. Enabled by the Unity Studies, the inclusion of these LMIC results increases the global representativeness of our review. This experience highlights the importance of open data practices and sharing early aggregate results for the collation and analysis of timely data, particularly during public health emergencies.^44^As vaccination coverage increases across the globe in high and upper-middle income countries, monitoring the transmission dynamics of COVID-19 in LMIC settings, where vaccination coverage is often still low, will be critical to overcome the pandemic.^45^

Our search strategy was highly sensitive and robust. We used broad search terms and language inclusion to capture as many relevant investigations as possible. However, we may have missed relevant investigations as we did not screen the reference lists of included investigations or other systematic reviews of hSAR. A further strength of this systematic review is that we attempted to clarify or confirm unclear or poorly reported items and contacted study investigators to request additional information.

In the future, we recommend focusing on the design of country-specific pre-planned (“at the ready”) standardised FFX and HHTIs with quality implementation by multi-disciplinary teams, preferably piloted in the field through ‘peace time’ exercises. These studies should be conducted in representative settings with available capacity, or, through collaborations to develop local capacity where it is otherwise limited. This coordinated approach would maximise opportunity to rapidly enact FFX and HHTIs during the early stages of the emergence of novel pathogens, especially respiratory viruses of pandemic potential like influenza and coronaviruses, allowing early characterisation of transmissibility and infection-severity to inform public health responses.^46^ Specific consideration should be given to prior ethics approval, governance, data collection methods and infrastructure, and resource requirements in advance of activation during a public health emergency.^47^ In addition to common protocols, tools to assist quality implementation (e.g. standard operating procedures for sample collection and publicly available data analysis scripts) and dissemination of results (e.g. scientific writing skill development) are required. Furthermore, although short-term assistance can support ad hoc investigations when required, initiatives to build and strengthen surveillance and laboratory capacity in LMICs – such as the WHO’s Pandemic Influenza Preparedness Framework – are a much more sustainable approach.^48,49^ Through long-term investment and development, LMICs can enhance surveillance and implement the operational research required to monitor co-circulating influenza and SARS-CoV-2 viruses in near-real-time, and detect the emergence of novel respiratory viruses of pandemic potential. In-country capacity building should be prioritised to ensure fit-for-purpose analytic methods for producing robust statistical inferences are implemented, such as those developed specifically for analysing household infection dynamics.^50^ We further recommend development of HHTI-specific guidelines and checklists for reporting, such as those developed for clinical trials^51^ or observational studies^52,53^, and those introduced in the HHTI critical appraisal tool.^17^

## Conclusion

As the COVID-19 pandemic progresses, FFX and HHTIs remain a critical tool to monitor population immunity to, and transmission dynamics and infection-severity of SARS-CoV-2, including the emergence of new genetic variants. These data are crucial to inform the ongoing response in different resource settings. Indeed, such estimates are key for regional and global modelling and forecasting to inform optimal application of PHSM and allocate pandemic resources including COVID-19 vaccines. The large and unexplained variance in hSAR estimates indicates the need for improved standardisation in the planning, conduct and analysis of HHTIs. Greater emphasis on clear and comprehensive reporting of HHTIs, and the context in which they are conducted, is required to facilitate more nuanced analysis of the sources of heterogeneity. High-quality FFX and HHTIs should continue to be conducted, ideally within a standardised framework such as the WHO Unity Studies initiative, and be supported to guide evidence-based pandemic preparedness and response efforts for SARS-CoV-2, influenza and future novel respiratory viruses.

## Data Availability

All data produced in the present work are contained in the manuscript as references (peer-reviewed publications) or at: https://zenodo.org/communities/unity-sero-2021/?page=1&size=20

## Unity Studies Collaborator Group

UNITY Studies Collaborator Group: Mikias A Alemu^15^, Yasir Alvi^16^, Elizabeth A Bukusi^17^, Pui Shan Chung^12^, Davaalkham Dambadarjaa^18^, Ayan K Das^19,20^, Timothée Dub^21^, Diba Dulacha^22^, Faiqa Ebrahim^23^, Maritza A González-Duarte^24^, Dinuka Guruge^25^, Damaris C Heredia-Melo^26^, Amy Herman-Roloff^27^, Belinda L Herring^2^, Farzana Islam^28^, Kamal Chandima Jeewandara^29^, Shashi Kant^30^, Richard Lako^31^, Juliana Leite^13^, Gathsaurie Neelika Malavige^32^, Undram Mandakh^33^, Warisha Mariam^34^, Tsogt Mend^35^, Valerie A Mize^22^, Sanjin Musa^36,37^, Hanna Nohynek^21^, Olushayo O Olu^22^, May B Osorio-Merchán^24^, Dmitriy Pereyaslov^1^, James Ransom^§38^, Lubna Al Ariqi^9^, Wasiq Khan^9^, Sonal Saxena^39^, Pragya Sharma^34^, Aswathy Sreedevi^40^, Mini Satheesh^41,42^, KJ Subhashini^30^, Beth A Tippet-Barr^26,43,^, Anjua Usha^44^, Joseph F Wamala^22^, Shambel H Watare^15^, Kapil Yadav^30^, Francis Y Inbanathan^11^

^§^deceased

^15^Ethiopian Public Health Institute

^16^Department of Community Medicine, Hamdard Institute of Medical Sciences and Research, New Delhi, India

^17^Kenya Medical Research Institute, Kenya

^18^ School of Public Health, Mongolian National University of Medical Sciences, Ulaanbaatar, Mongolia

^19^Department of Microbiology, Hamdard Institute of Medical Science and Research, New Delhi, India

^20^Hakeem Abdul Hameed Centenary Hospital, New Delhi, India

^21^Department of Health Security, Finnish Institute for Health and Welfare, Helsinki, Finland

^22^WHO Country Office, Juba, South Sudan

^23^World Health Organization Country Office, Ethiopia

^24^National Institute of Health, Colombia

^25^Colombo Municipality Council, Colombo, Sri Lanka

^26^U.S. Centers for Disease Control and Prevention, Kenya

^27^Department of Community Medicine, Hamdard Institute of Medical Sciences and Research, New Delhi, India

^28^Hamdard Institute of Medical Sciences & Research (HIMSR), New Delhi, India

^29^Allergy Immunology and Cell Biology Unit, Department of Immunology and Molecular Medicine, Faculty of Medical Sciences, University of Sri Jayewardenepura, Gangodawila, Nugegoda, Sri Lanka

^30^Centre for Community Medicine, All India Institute of Medical Sciences, New Delhi, India

^31^Ministry of Health, South Sudan

^32^Faculty of Medical Sciences, University of Sri Jayawardenapura, Nugegoda, Sri Lanka

^33^Mongolian National University of Medical Sciences, Ulaanbaatar, Mongolia

^34^Department of Community Medicine, Maulana Azad Medical College, New Delhi, India

^35^National Center for Communicable Diseases, Ulaanbaatar, Mongolia

^36^Institute for Public Health of the Federation of Bosnia and Herzegovina, Sarajevo, Bosnia and Herzegovina

^37^Sarajevo School of Science and Technology, Sarajevo, Bosnia and Herzegovina

^38^Centers for Disease Control and Prevention, South Sudan

^39^Department of Microbiology, Maulana Azad Medical College, New Delhi, India

^40^Department of Community Medicine, Amrita Institute of Medical Sciences, Kochi, Kerala, India

^41^Kerala University of Health Sciences, Kerala, India

^42^Government Medical College, Thiruvananthapuram, Kerala, India

^43^Nyanja Health Research Institute, Salima, Malawi

^44^Regional Prevention of Epidemic and Infectious Disease cell, Government of Kerala, Kerala, India

## Acknowledgements

Thank you to Miu-Ling Wong, Hong Kong University, for her assistance with translation of articles in Mandarin.

We thank colleagues: Mike Ryan, Ketevan Glonti, Aisling Vaughan, Tony Wurda, Johson Muki, Yona Kenyi Manoah, Manuela Alphose Juma, Suzy Agrey Abas, Musah Kwaku Bukaru, Adamu Tayachew, Negussie Yohannes, Zewdu Assefa, Mengistu Biru, Musa Emmanuel, Margaret Mburu, Jayne Lewis-Kulzer, Pamela Murnane, Rachael Joseph, Elizabeth Oele, John Ndyahikayo, Ayesheshem A Tegegne, Arash Rashidian, Anna Solastie, Anu Haveri, Niina Ikonen, Lotta Hagberg, Oona Liedes, Mia Blazevic, Anes Joguncic, Faris Dizdar, Palo Mirza, Franklyn E Prieto-Alvarado, Ruth C Figueroa-Baez, Susanne C Ardila-Roldan, Mohammad Ahmad, Anisur Rahman, Shivani Rao, Mongjam Meghachandra Singh, Saurav Basu, Vikas Manchanda, Rohit Chawla, Faheem Ahmed, Mridu Dudeja, Farishta HD Singh, Iqbal Alam, Sushovan Roy, Ekta Gupta, Anil Kumar, Neethu Mohan, Soumya Gopakumar, Geetha Raveendranath, Deshni Jayathikala, Inoka S Aberathna, Saubhagya Danasekara, Buddini Gunathilake, Ruwan Wijayamuni, Ambaselmaa Amarjarga, Bilegt Altangerel, Temuulen Enebish, Oyunsuren Enebish, Ariuntuya Ochirpurev.

We would especially like to thank all WHO UNITY Study Collaborator Group, who are also named co-authors of this paper. These and other collaborators are recognized on a dedicated webpage on the UNITY and WHO website^1^ in all the countries who embarked in this global response effort to COVID-19, as well as all individuals who supported, conducted or participated in each of the studies supported.

The authors alone are responsible for the views expressed in this article and they do not necessarily represent the views, decisions, or policies of the institutions with which they are affiliated.

## Data sharing

The data extraction template, data extraction forms, data used for analysis and the analysis code are publicly available on Zenodo in the WHO Unity Studies Global SARS-CoV-2 Seroepidemiological Investigations community.^2^

### Appendix

#### Detailed searches

##### 1. Medline – searched 17^th^ August 2021

COVID-19/ or (COVID-19 or COVID19 or SARS-CoV-2 or coronavirus).tw AND

(Household transmission OR secondary attack rate* OR close contact* OR contact transmission OR contact attack rate* OR family transmission).tw

##### 2. EMBASE – searched 17^th^ August 2021

coronavirus disease 2019/ or (COVID-19 or COVID19 or SARS-CoV-2 or coronavirus).tw AND

(Household transmission OR secondary attack rate* OR close contact* OR contact transmission OR contact attack rate* OR family transmission).tw

##### 3. Web of Science and Scopus – searched 17^th^ August 2021

COVID-19 or COVID19 or SARS-CoV-2 or coronavirus

AND

“household transmission” OR “secondary attack rate*” OR “close contact*” OR “contact transmission” OR “contact attack rate*” OR “family transmission”

##### 4. medRxiv and bioRxiv (through medRxiv – COVID-19 papers have been cross-indexed) – searched 30^th^ August 2021

Advanced Search of (abstracts or titles)

Covid “household transmission”

Covid “secondary attack rate”

Covid “close contact”

Covid “contact transmission”

Covid “contact attack rate”

Covid “family transmission

#### Detailed eligibility criteria

##### Inclusion criteria

For screening of abstracts and full-text:

- Any (published) investigation/study involving five or more discrete households with at least one laboratory confirmed index case of COVID-19 and all their household contacts or WHO supported investigations
- Investigations will need to provide information on transmission dynamics, severity and clinical spectrum of COVID-19 within the household setting. Specifically, they need to report on the household secondary attack rate (infection or clinical) with a measure of uncertainty, or contain sufficient data to calculate the secondary attack rate (i.e., number of household contacts that become cases, total number of household contacts and household sizes) and may also report on the following: severity indicators such as the asymptomatic proportion of cases, hospitalisation and ICU admission rate, case fatality rate and transmission parameters such as the serial interval.
- Eligible papers will be restricted to the following languages: English, Chinese, French, Russian and Spanish, Portuguese, German and Italian.

For full-text only:

- These studies will need to be aligned to the WHO Unity Studies FFX and HH transmission investigation protocols (available at: https://www.who.int/emergencies/diseases/novel-coronavirus-2019/technical-guidance/early-investigations)
  - household recruitment is triggered by detection of a RT-PCR-confirmed index case
  - longitudinally collected (prospective or retrospective) data from all household members including epidemiological/virological or serological data (within the 28-day period from symptom onset in the primary case or household enrolment)
  - Active follow-up of households (through case investigation and contact tracing)

##### Exclusion criteria

- Duplicate studies will be removed. Where multiple papers are published on the same cohort (i.e., interim and final findings, subgroup analyses and secondary data analyses), we will use the most complete and relevant data from the article where summary statistics are appropriately reported.
- Studies that don’t report sufficient information (i.e., conference abstracts or where full-text is not available or where additional information can’t be accessed upon contacting study investigators).

We investigated if the estimates varied by the following subgroups: country/region where study was conducted, pandemic course (epidemic activity), income setting, predominantly circulating variant at the time of the investigation/study was conducted, size of household, source population, implementation methods and index case symptom status. If data were available, we investigated if the estimates varied by sex, and age groups of the index case. We also plan to investigate if the estimates vary by publication status (i.e., comparing investigations in the literature to unpublished investigations).

#### Changes from PROSPERO registration and other details

- Adapted criteria for WHO Unity Studies investigations to be included irrespective of number of households included and follow-up of all household contacts. This allowed us to consider inclusion of estimates that would otherwise not be included in line with our objectives. Note that all included WHO-supported investigations had at least five households.
- We did not screen reference lists of included articles. Unpublished articles and reports provided from WHO often did not contain a reference list, and we believe that our search strategy was very comprehensive and sensitive.
- Changes to subgroup analyses were determined prior to the conduct of analysis (Initial plans were as follows: We investigated if the estimates varied by the following subgroups: country/region where study was conducted, pandemic course (epidemic activity), income setting, predominantly circulating variant at the time of the investigation/study was conducted, size of household, source population, implementation methods and index case symptom status. If data were available, we investigated if the estimates varied by sex, and age groups of the index case. We also plan to investigate if the estimates vary by publication status)
- Authors in the collaborator group were included based on the ICJME criteria. Authorship was not offered in exchange for contribution of early data to the review.
- We initially intended to review all transmission and severity parameters. We decided to focus on the hSAR and hSCAR only to enable more detailed analysis of collected data.
- Description of the statistical analysis was modified to reflect change in meta-analysis models used in the analysis.
- Due to the significant amount of observed heterogeneity, we did not present a pooled HSAR and hSCAR estimate and did not visually assess funnel plots of effect size versus standard error and used Egger’s test to evaluate bias due to small study effect.(18)
- Additionally, we planned to conduct a sensitivity analysis by fitting a beta-binomial model as applied in Thompson et al. to explore whether the chosen model impacted the results. This was not undertaken as we decided to not produce a pooled estimate.

#### Detailed changes from the planned statistical analyses

| Planned approach | Updated approach | Reason for change |
| --- | --- | --- |
| Meta-analyses would be undertaken to produce pooled hSAR and hSCAR estimates. | No pooled estimates of hSAR or hSCAR were produced. A 95% prediction interval was produced. | There was substantial heterogeneity in estimated hSAR. We anticipated that we may be able to explain these sources of heterogeneity through several pre-specified subgroup analyses and provide pooled estimates at a finer resolution (i.e., by subgroup).<br>The primary meta-analysis produced a high $I^2$ value of 99.7%, indicating a high degree of heterogeneity as anticipated. The high degree of heterogeneity was also reflected in the wide range of reported hSAR estimates (2% – 90%) and prediction interval (3% – 71%). Substantial heterogeneity was still present within these pre-specified subgroups, as well as a number of post-hoc subgroups.<br>Given our understanding of the differences in the source of the estimated hSAR in each investigation, and the residual heterogeneity that was present upon meta-analysis within subgroups, it was decided it was not appropriate to report |
|  |  | any pooled estimates. Instead, we added in the estimated 95% prediction interval to help illustrate the large variation in results. |
| A beta-binomial mixed-effects model would be fit for primary meta-analysis and a binomial-normal model would be fit for sensitivity analysis. | A binomial-normal mixed-effects meta-analysis model was used for the primary meta-analysis. | Both methodologies have been demonstrated to be appropriate for the chosen outcome measures <sup>24,28</sup>, and we intended to utilise the beta-binomial model for the primary analysis and the binomial-normal sensitivity analysis.<br>The binomial-normal model had existing implementations in our chosen statistical package <sup>31</sup>, and so we decided to utilise this as for the primary analysis. No sensitivity analysis was undertaken due to the decision not to pool hSAR or hSCAR estimates, as described above. |
| We would assess reporting biases through 1) visual inspection of funnel plots of the effect size, and; 2) using Egger's test to evaluate bias due to small study effect. | No assessment of reporting biases was made. | Because we chose not to produce pooled estimates any of the outcomes of interest, there was no need to assess the impact of reporting biases on these pooled estimates. |
| Pre-specified subgroups: 1) Income setting; 2) Predominantly circulating variant at the time of investigation; 3) Secondary case ascertainment methods and; 4) Methodological quality of investigation. | Additional post-hoc subgroups: 1) Duration of follow-up of household contacts; 2) Stricter adherence to the Unity protocol, and; 3) Use of serological testing to ascertain secondary cases. | The high heterogeneity between hSAR estimates ($I^2$ value of 99.7%) was not reduced in pre-specified subgroup analyses ($I^2$ values ranging between 99.4% – 99.6%).<br>To address this, we defined three additional post-hoc subgroups we anticipated may account for heterogeneity. |

#### Detailed questions used for methodological quality assessment

The questions detailed in the HHTI critical appraisal tool (13) used to assess these studies are as follows:

1. Was the study planning, recruitment and data collection appropriately timed to achieve the objectives of the investigation?
2. Was the cohort of households enrolled as a consequence of an adequate method of case ascertainment?
3. Was a definition of “household” provided?
4. Were all eligible cases and all householders enrolled into the investigation? 5.
5. 
  a. Were subsequent cases identified and ascertained using appropriate methods to calculate the hSAR?
  b. Were subsequent cases identified and ascertained using appropriate methods to calculate the hSCAR?
6. 
  a. Were households followed up for a period sufficient to measure primary outcomes?
  b. Did all primary cases and contacts remain part of the “household” for the duration of the investigation?
7. 
  a. Were steps taken to ensure that householders were susceptible at the time of enrolment?
  b. Were steps taken to ensure that subsequent infections were due to exposure within the household?
8. Are the methods for analysis appropriate given the study context and design, participant definitions and the measurement of outcomes?
9. Has loss to follow-up been appropriately accounted for in the estimated outcomes?
10. Has any missing data been appropriately accounted for in the estimated outcomes?

