## Supplementary Material for "Transmission of SARS-CoV-2 in standardised First Few X cases and household transmission investigations: a systematic review and meta-analysis"

**Supplementary Table 1.** Detailed descriptions and characteristics of investigations included in the systematic review.

| Author (ref) | Country | WHO Region | Income Status | HRP Status | No. Index Cases | No. Households | No. Household Contacts, Included/Total | No. Secondary Cases | Timing of Recruitment | Household Study Design | Secondary Case Ascertainment Methods(s) | Data Collection Method | Duration of Follow Up |
|---|---|---|---|---|---|---|---|---|---|---|---|---|---|
| **Angulo-Bazan, Y. et al.[1]** | Peru | AMR | Upper-Middle | Yes | 52 | 52 | 236 | 125 [serology]; 162 [symptoms] | 23 April 2020 - 2 May 2020 | Yes | Serology [hSAR]; symptoms only [hSCAR] | Retrospective | Unclear |
| **Areekal, B. et al.[2]** | India | SEAR | Lower-Middle | No | 212 | 212 | 849 | 221 [lab diagnosis]; 132 [symptoms] | June 2020 - July 2020 | No | Unclear [hSAR]; symptoms only [hSCAR] | Prospective | 14 days or less |
| **Bi, Q. et al. [3]** | China | WPR | Upper-Middle | No | 244 | Unknown | 686 | 77 | 14 January 2020 - 12 February 2020 | No | RT-PCR | Retrospective | 14 days or less |
| **Boscolo-Rizzo, P. et al.[4]** | Italy | EUR | High | No | 179 | 179 | 121 [hSAR]; 296 [hSCAR] | 54 [RT-PCR]; 233 [symptoms] | March 2020 - April 2020 | Yes | RT-PCR [hSAR]; symptoms only [hSCAR] | Retrospective | Unclear |
| **Buchan, S.A. et al.[5]** | Canada | AMR | High | No | 5617 | 5617 | 15597 | 3397 | 7 February 2021 - 27 February 2021 | Yes | Unclear | Prospective | 14 days or less |
| **Burke, R.M. et al. (A)[6]** | United States of America | AMR | High | No | 69 | 69 | 193/201 | 18 [RT-PCR]; 62 [symptoms] | January 2020 - April 2020 | Yes | RT-PCR [hSAR]; symptoms only [hSCAR] | Prospective | 14 days or less |
| **Burke, R.M. et al. (B)[7]** | United States of America | AMR | High | No | 9 | 9 | 15 | 2 [RT-PCR]; 8 [symptoms] | January 2020 | No | RT-PCR [hSAR]; symptoms only [hSCAR] | Prospective | 14 days or less |
| **Carazo, S. et al.[8]** | Canada | AMR | High | No | 3823 | 3823 | 9096 | 2718 | 1 March 2020 - 14 June 2020 | No | Symptoms | Retrospective | 14 days or less |
| **Cerami, C. et al.[9]** | United States of America | AMR | High | No | 100 | 100 | 176/182 | 106 [RT-PCR or serology]; 91 [symptoms] | April 2020 - October 2020 | Yes | RT-PCR or serology [hSAR]; symptoms only [hSCAR] | Prospective | Greater than 14 days |
| **Chaw, L. et al.[10]** | Brunei Darussalam | WPR | High | No | 19 | 19 | 123 | 16 | March 2020 | No | RT-PCR | Prospective | 14 days or less |
| **Cheng, H.Y. et al.[11]** | Taiwan, China | WPR | High | No | 100 | 100 | 151 | 10 [RT-PCR]; 7 [symptoms] | 15 January 2020 - 18 March 2020 | No | RT-PCR [hSAR]; symptoms only [hSCAR] | Prospective | 14 days or less |
| **Dambadarjaa, D. et al.[12]** | Mongolia | WPR | Lower-Middle | No | 97 | 97 | 325 | 60 | November 2020 | No | RT-PCR | Prospective | 14 days or less |
| **Dattner, I. et al.[13]** | Israel | EUR | High | No | 637 | 637 | 3353 | 1510 [RT-PCR]; 1243 [symptoms] | 17 March 2020 - 3 May 2020 | Yes | RT-PCR [hSAR]; symptoms only [hSCAR] | Retrospective | Unclear |
| **Gomaa, M.R. et al.[14]** | Egypt | EMR | Lower-Middle | Yes | 23 | 23 | 98 | 88 [RT-PCR or serology]; 52 [symptoms] | April 2020 - October 2020 | Yes | RT-PCR or serology [hSAR]; symptoms only [hSCAR] | Prospective | 14 days or less |
| **Grijalva, C.G. et al.[15]** | United States of America | AMR | High | No | 101 | 101 | 137/191 | 48 | April 2020 - September 2020 | Yes | RT-PCR | Prospective | 14 days or less |
| **Heredia-Melo, D.C. et al.[16]** | Colombia | AMR | Upper-Middle | Yes | 99 | 99 | 315/334 | 140 | September 2020 - January 2021 | No | RT-PCR and symptoms | Prospective | 14 days or less |
| **Hsu, C.Y. et al.[17]** | Taiwan, China | WPR | High | No | 18 | 26 | 102 | 47 | 28 January 2020 - 28 February 2021 | Yes | RT-PCR | Retrospective | 14 days or less |
| **Hu, P. et al.[18]** | China | WPR | Upper-Middle | No | 82 | 82 | 267 | 46 | Before 5 March 2020 | No | RT-PCR | Retrospective | 14 days or less |
| **Hu, S.X. et al.[19]** | China | WPR | Upper-Middle | No | 284 | 115 | 1021 | 99 | 23 January 2020 - 2 April 2020 | No | RT-PCR | Prospective | 14 days or less |
| **Islam, F. et al.[20]** | India | SEAR | Lower-Middle | No | 99 | 99 | 254/318 | 141 | 15 December 2020 - 16 June 2021 | Yes | RT-PCR | Prospective | Greater than 14 days |
| **Jashaninejad, R. et al.[21]** | Iran (Islamic Republic of) | EMR | Lower-Middle | Yes | 323 | 323 | 989 | 314 | May 2020 - July 2020 | No | RT-PCR | Prospective | 14 days or less |
| **Jeewandara, C. et al.[22]** | Sri Lanka | SEAR | Lower-Middle | No | 39 | 39 | 1093 | 85 | 15 April 2020 - 19 May 2020 | No | RT-PCR | Prospective | 14 days or less |

| Author (ref) | Country | WHO Region | Income Status | HRP Status | No. Index Cases | No. Households | No. Household Contacts, Included/Total | No. Secondary Cases | Timing of Recruitment | Household Study Design | Secondary Case Ascertainment Methods(s) | Data Collection Method | Duration of Follow Up |
|---|---|---|---|---|---|---|---|---|---|---|---|---|---|
| **Jeong, E.K. et al.[23]** | Republic of Korea | WPR | High | No | 30 | 30 | 119 | 9 | 24 January 2020 - 10 March 2020 | No | Symptoms | Retrospective | 14 days or less |
| **Jing, Q.L. et al.[24]** | China | WPR | Upper-Middle | No | 159 | 159 | 529 | 93 | 7 January 2020 - 18 February 2020 | Yes | RT-PCR | Retrospective | 14 days or less |
| **Korean CDC[25]** | Republic of Korea | WPR | High | No | 9 | 9 | 119 | 9 | 24 January 2020 - 10 March 2020 | No | Unclear | Prospective | 14 days or less |
| **Koureas, M. et al.[26]** | Greece | EUR | High | No | 30 | 30 | 164 | 62 | 8 April 2020 - 4 June 2020 | No | RT-PCR | Prospective | 14 days or less |
| **Kuba, Y. et al.[27]** | Japan | WPR | High | No | 78 | 78 | 174 | 21 [RT-PCR]; 21 [symptoms] | 14 February 2020 - 31 May 2020 | Yes | RT-PCR and symptoms [hSAR]; symptoms only [hSCAR] | Prospective | 14 days or less |
| **Kuwelker, K. et al.[28]** | Norway | EUR | High | No | 112 | 112 | 179/245 | 85 [RT-PCR or serology], 130 [symptoms] | 28 February 2020 - 4 April 2020 | Yes | RT-PCR or serology [hSAR]; symptoms only [hSCAR] | Prospective | Greater than 14 days |
| **Laxminarayan, R. et al.[29]** | India | SEAR | Lower-Middle | No | 998 | 998 | 4065 | 380 | 5 March 2020 - 1 August 2020 | No | RT-PCR | Retrospective | 14 days or less |
| **Layan, M. et al.[30]** | Israel | EUR | High | No | 215 | 210 | 687 | 269 | December 2020 - April 2021 | Yes | RT-PCR | Prospective | 14 days or less |
| **Lewis, N.M. et al.[31]** | United States of America | AMR | High | No | 58 | 58 | 188/197 | 52 [RT-PCR or serology]; 136 [symptoms] | 22 March 2020 - 25 April 2020 | Yes | RT-PCR or serology [hSAR]; symptoms only [hSCAR] | Prospective | 14 days or less |
| **Li, F. et al.[32]** | China | WPR | Upper-Middle | No | 24985 | 24985 | 52822 | 8447 | 2 December 2019 - 18 April 2020 | Yes | Unclear | Retrospective | 14 days or less |
| **Li, W. et al.[33]** | China | WPR | Upper-Middle | No | 105 | 105 | 392 | 64 [RT-PCR]; 55 [symptoms] | 1 January 2020 - 20 February 2020 | Yes | RT-PCR [hSAR]; symptoms only [hSCAR] | Prospective | 14 days or less |
| **Liu, T. et al.[34]** | China | WPR | Upper-Middle | No | 1206 | 1206 | 4707 | 410 | 10 January 2020 - 15 March 2020 | No | RT-PCR | Retrospective | 14 days or less |
| **Loenenbach, A. et al.[35]** | Germany | EUR | High | No | 38 | 38 | 92 | 34 | January 2021 - February 2021 | No | RT-PCR and symptoms | Prospective | 14 days or less |
| **Luo, L. et al.[36]** | China | WPR | Upper-Middle | No | 197 | 197 | 1015 | 105 | 13 January 2020 - 6 March 2020 | No | RT-PCR | Prospective | 14 days or less |
| **Maltezou, H.C. et al.[37]** | Greece | EUR | High | No | 133 | 133 | 846 | 295 | 26 February 2020 - 30 June 2020 | No | RT-PCR and symptoms | Retrospective | 14 days or less |
| **Marcato, A.J. et al.[38]** | Australia | WPR | High | No | 101 | 96 | 286/300 | 40 | April 2020 - October 2020 | Yes | RT-PCR | Prospective | 14 days or less |
| **Marks, M. et al.[39]** | Spain | EUR | High | No | 282 | 282 | 382 | 91 | 17 March 2020 - 28 April 2020 | No | RT-PCR | Prospective | 14 days or less |
| **Miller, E. et al.[40]** | United Kingdom | EUR | High | No | 117 | 117 | 248 | 75 | 30 March 2020 - 17 November 2020 | Yes | RT-PCR or serology | Prospective | 14 days or less |
| **Miyahara, R. et al.[41]** | Japan | WPR | High | No | 306 | 306 | 775 | 147 | 22 February 2020 - 31 May 2020 | Yes | RT-PCR | Retrospective | Greater than 14 days |
| **Mize, V. et al.[42]** | South Sudan | AFR | Low | Yes | 29 | 29 | 13089 | 1 | 8 June 2020 - 3 December 2020 | No | RT-PCR or serology | Prospective | Greater than 14 days |
| **Musa, S. et al.[43]** | Bosnia and Herzegovina | EUR | Upper-Middle | No | 383 | 360 | 747 | 119 [RT-PCR and symptoms]; 103 [symptoms] | 3 August 2020 - 23 December 2020 | Yes | RT-PCR and symptoms [hSAR]; symptoms only [hSCAR] | Prospective | 14 days or less |
| **Ng, O.T. et al.[44]** | Singapore | WPR | High | No | 581 | 578 | 1779/1863 | 134 [RT-PCR or serology]; 468 [symptoms] | 23 January 2020 - 3 April 2020 | Yes | RT-PCR or serology [hSAR]; symptoms only [hSCAR] | Retrospective | 14 days or less |
| **Nohynek, H. et al.[45]** | Finland | EUR | High | No | 37 | 37 | 87/90 | 42 | Mar-20 | Yes | RT-PCR or serology | Prospective | Unclear |
| **Park, S.Y., et al.[46]** | Republic of Korea | WPR | High | No | 97 | Unknown | 225 | 34 | 21 February 2020 - 8 March 2020 | No | RT-PCR | Prospective | 14 days or less |
| **Pett, J. et al.[47]** | United Kingdom | EUR | High | No | 27 | 27 | 44 | 7 [RT-PCR]; 7 [symptoms] | 26 February 2020 - 26 April 2020 | No | RT-PCR and symptoms [hSAR]; symptoms only [hSCAR] | Prospective | 14 days or less |

| Author (ref) | Country | WHO Region | Income Status | HRP Status | No. Index Cases | No. Households | No. Household Contacts, Included/Total | No. Secondary Cases | Timing of Recruitment | Household Study Design | Secondary Case Ascertainment Methods(s) | Data Collection Method | Duration of Follow Up |
|---|---|---|---|---|---|---|---|---|---|---|---|---|---|
| **Phucharoen, C. et al.**[48] | Thailand | SEAR | Upper-Middle | No | 77 | 63 | 171 | 82 | 20 March 2020 - 2 May 2020 | No | RT-PCR | Prospective | Unclear |
| **Pung, R. et al.**[49] | Singapore | WPR | High | No | 265 | 265 | 875 | 33 | Before 21 March 2020 | Yes | RT-PCR and symptoms | Prospective | 14 days or less |
| **Ransom, J. et al.**[50] | South Sudan | AFR | Low | Yes | 26 | 26 | 30 [hSAR]; 42 [hSCAR] | 17 [serology]; 2 [symptoms] | Jun-20 | Yes | Serology [hSAR]; symptoms only [hSCAR] | Prospective | Unclear |
| **Ratovoson, R. et al.**[51] | Madagascar | AFR | Low | No | 33 | 33 | 179/192 | 53 | 19 March 2020 - 30 July 2020 | Yes | RT-PCR or serology | Prospective | Greater than 14 days |
| **Reukers, D.F.M. et al.**[52] | Netherlands | EUR | High | No | 55 | 55 | 187 | 78 [RT-PCR or serology]; 79 [symptoms] | 24 March 2020 - 6 April 2020 | Yes | RT-PCR or serology [hSAR]; symptoms only [hSCAR] | Prospective | Greater than 14 days |
| **Ripabelli, G. et al.**[53] | Italy | EUR | High | No | Unknown | 34 | 59 | 16 | May-20 | No | RT-PCR | Retrospective | Greater than 14 days |
| **Salihefendic, N. et al.**[54] | Bosnia and Herzegovina | EUR | Upper-Middle | No | 25 | 25 | 123 | 76 | March 2020 - December 2020 | Yes | RT-PCR or serology | Prospective | Unclear |
| **Semakula, M. et al.**[55] | Rwanda | AFR | Low | Yes | 528 | 153 | 615 | 18 | 14 March 2020 - 20 July 2020 | No | RT-PCR | Retrospective | 14 days or less |
| **Shah, K. et al. (A)**[56] | India | SEAR | Lower-Middle | No | 72 | 72 | 287 | 5 | March 2020 - July 2020 | Yes | Unclear | Prospective | 14 days or less |
| **Shah, K. et al. (B)**[57] | India | SEAR | Lower-Middle | No | 74 | 74 | 386 | 34 | March 2020 - July 2020 | Yes | Unclear | Unclear | Greater than 14 days |
| **Shambel, H. et al.**[58] | Ethiopia | AFR | Low | Yes | 100 | 100 | 221/300 | 100 | 19 May 2020 - 15 June 2020 | No | RT-PCR | Prospective | 14 days or less |
| **Sharma, P. et al.**[59] | India | SEAR | Lower-Middle | No | 146 | 146 | 202/270 | 67 [RT-PCR]; 28 [symptoms] | 28 December 2020 - 28 June 2021 | Yes | RT-PCR [hSAR]; symptoms only [hSCAR] | Prospective | Greater than 14 days |
| **Son, H. et al.**[60] | Republic of Korea | WPR | High | No | 108 | Unknown | 196 | 16 | 21 February 2020 - 24 March 2020 | No | RT-PCR | Prospective | 14 days or less |
| **Soriano-Arandes, A. et al.**[61] | Spain | EUR | High | No | 270 | 341 | 3392 | 2091 [RT-PCR]; 1386 [symptoms] | 1 July 2020 - October 2020 | Yes | RT-PCR [hSAR]; symptoms only [hSCAR] | Prospective | Unclear |
| **Sreedevi, A. et al.**[62] | India | SEAR | Lower-Middle | No | 147 | 147 | 364 | 159 [RT-PCR]; 94 [symptoms] | 21 December 2020 - 30 July 2021 | Yes | RT-PCR [hSAR]; symptoms only [hSCAR] | Prospective | Greater than 14 days |
| **Sun, W.W. et al.**[63] | China | WPR | Upper-Middle | No | 149 | 149 | 697 | 240 | 8 January 2020 - 6 February 2020 | No | RT-PCR | Retrospective | 14 days or less |
| **Teherani, M.F. et al.**[64] | United States of America | AMR | High | No | 32 | 32 | 144 | 67 | 16 March 2020 - 14 June 2020 | Yes | Symptoms | Prospective | 14 days or less |
| **Telle, K. et al.**[65] | Norway | EUR | High | No | 7548 | 7548 | 19443 | 4613 | 1 March 2020 - 1 January 2021 | No | RT-PCR | Retrospective | 14 days or less |
| **Thiel, S.L. et al.**[66] | Liechtenstein | EUR | High | No | 81 | 81 | 109/127 | 35 [serology]; 49 [symptoms] | March 2020 - April 2020 | No | Serology [hSAR]; symptoms only [hSCAR] | Prospective | 14 days or less |
| **Tibebu, S. et al.**[67] | Canada | AMR | High | No | 29352 | 29352 | 84125 | 16404 | July 2020 - November 2020 | Yes | Symptoms | Prospective | Greater than 14 days |
| **Tippett Barr, B. et al.**[68] | Kenya | AFR | Lower-Middle | Yes | 125 | 51 | 156 | 24 | June 2020 - October 2020 | No | RT-PCR | Prospective | 14 days or less |
| **Walker, J.G. et al.**[69] | United States of America | AMR | High | No | 917 | 917 | 1766 | 655 | February 2020 - June 2020 | No | RT-PCR | Prospective | 14 days or less |
| **Wang, Y. et al.**[70] | China | WPR | Upper-Middle | No | 124 | 41 | 335 | 77 | 28 February 2020 - 27 March 2020 | Yes | Unclear | Prospective | 14 days or less |
| **Wang, Z. et al.**[71] | China | WPR | Upper-Middle | No | 78 | 78 | 155 | 47 [RT-PCR]; 104 [symptoms] | 13 February 2020 - 14 February 2020 | Yes | RT-PCR [hSAR]; symptoms only [hSCAR] | Retrospective | 14 days or less |
| **Wei, L. et al.**[72] | China | WPR | Upper-Middle | No | 39 | 23 | 66 | 21 | January 2020 - February 2020 | Yes | Unclear | Unclear | 14 days or less |
| **Wilkinson, K. et al.**[73] | Canada | AMR | High | No | 102 | 102 | 279 | 41 [RT-PCR]; 41 [symptoms] | March 2020 - 28 April 2020 | Yes | RT-PCR and symptoms [hSAR]; symptoms only [hSCAR] | Prospective | 14 days or less |
| **Wu, J. et al.**[74] | China | WPR | Upper-Middle | No | 35 | 35 | 148 [hSAR]; 143 [hSCAR] | 48 [RT-PCR]; 51 [symptoms] | 17 January 2020 - 29 February 2020 | Yes | RT-PCR [hSAR]; symptoms only [hSCAR] | Prospective | Greater than 14 days |

| Author (ref) | Country | WHO Region | Income Status | HRP Status | No. Index Cases | No. Households | No. Household Contacts, Included/Total | No. Secondary Cases | Timing of Recruitment | Household Study Design | Secondary Case Ascertainment Methods(s) | Data Collection Method | Duration of Follow Up |
|---|---|---|---|---|---|---|---|---|---|---|---|---|---|
| **Wu, P. et al.[75]** | China | WPR | Upper-Middle | No | 393 | 393 | 1516 | 104 | 5 January 2020 - 7 April 2020 | No | RT-PCR | Retrospective | 14 days or less |
| **Wu, Y. et al.[76]** | China | WPR | Upper-Middle | No | 144 | Unknown | 280 | 50 | 23 January 2020 - 28 February 2020 | No | Unclear | Retrospective | 14 days or less |
| **Xie, W. et al.[77]** | China | WPR | Upper-Middle | No | 20 | 20 | 79 | 53 [RT-PCR]; 49 [symptoms] | 21 January 2020 - 14 February 2020 | Yes | RT-PCR [hSAR]; symptoms only [hSCAR] | Prospective | 14 days or less |
| **Yadav, K. et al.[78]** | India | SEAR | Lower-Middle | No | 148 | 148 | 645 | 197 [RT-PCR or serology]; 60 [symptoms] | 16 December 2020 - 24 June 2021 | Yes | RT-PCR or serology [hSAR]; symptoms only [hSCAR] | Prospective | Greater than 14 days |
| **Yi, B. et al.[79]** | China | WPR | Upper-Middle | No | 214 | 214 | 475 | 204 [RT-PCR and symptoms]; 224 [symptoms] | 22 January 2020 - 4 February 2020 | Yes | RT-PCR and symptoms [hSAR]; symptoms only [hSCAR] | Retrospective | 14 days or less |
| **Yi, C. et al.[80]** | China | WPR | Upper-Middle | No | 157 | 157 | 279 | 37 [RT-PCR]; 27 [symptoms] | January 2020 - March 2020 | No | RT-PCR [hSAR]; symptoms only [hSCAR] | Prospective | 14 days or less |

Abbreviations: ref, Reference; WHO, World Health Organization; HRP, Humanitarian Response Plan; AFR, Africa; AMR, Americas; EMR, Eastern Mediterranean; EUR, Europe; SEAR, South East Asia; WPR, Western Pacific; hSAR, household secondary infection attack rate; hSCAR, household secondary clinical attack rate; RT-PCR, reverse transcription polymerase chain reaction.

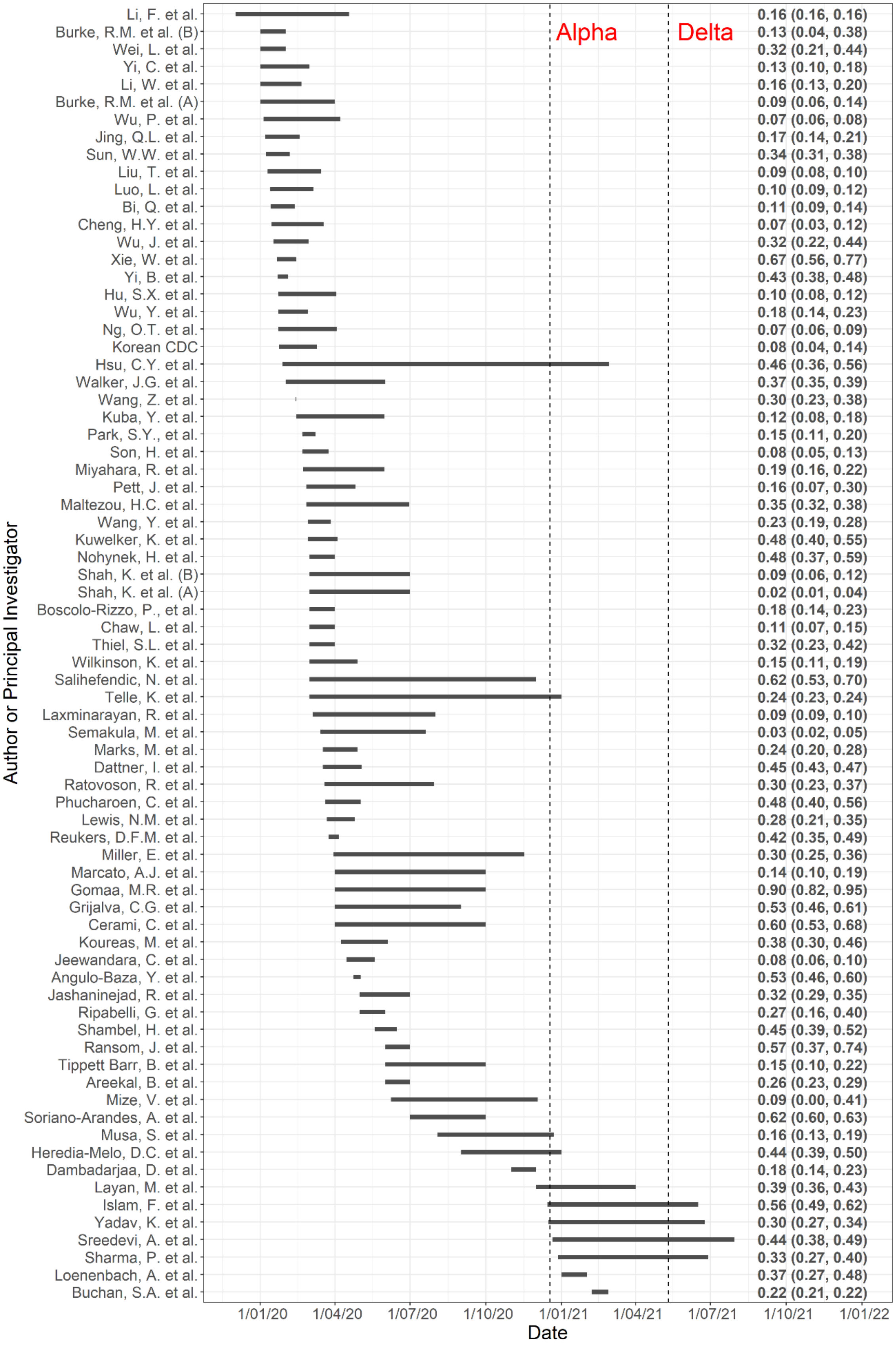


**Supplementary Figure 1**: Horizontal lines showing the approximate timing of investigations reporting a household secondary infection attack rate (hSAR) by month and year. Two of the 76 investigations reporting a hSAR are not plotted as the start date was not reported. Vertical lines represent the time at which variants Alpha and Delta were designated as Variants of Concern (VoC) by the WHO (18 December 2020, and 5 May 2021, respectively). Investigations are ordered by start dates. The hSAR and 95% confidence intervals are provided on the right-hand side.

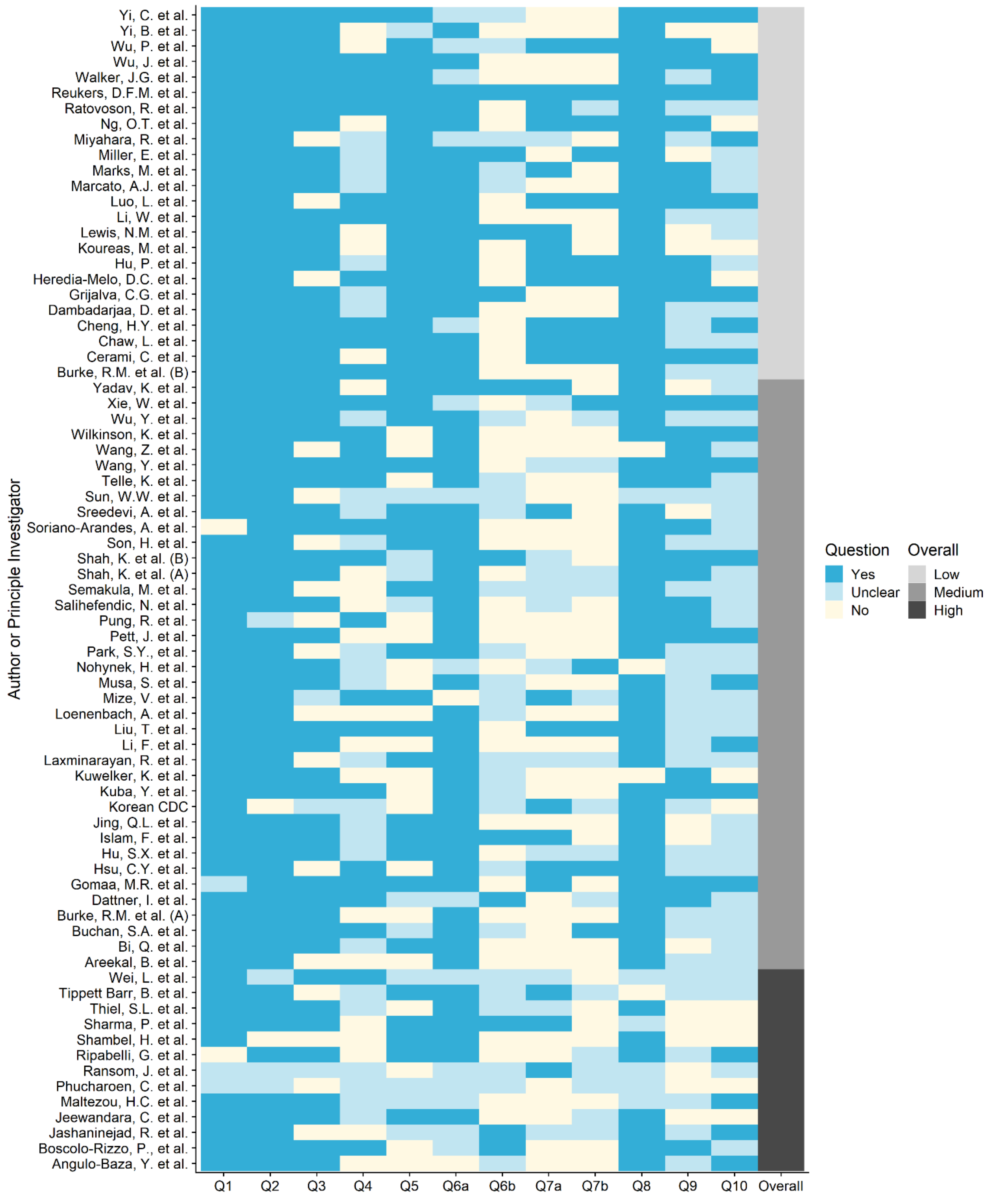


**Supplementary Figure 2.** Results of the critical appraisal tool as applied to investigations that reported household secondary infection attack rate (hSAR). Colours for Questions 1–10 indicate whether each was addressed in the investigation (dark blue) or not (cream), or instances where there was insufficient detail available to assess (light blue). An overall rating of the risk of bias is provided in the far-right column, with investigations rated Low (light grey), Medium (medium grey) or High (dark grey).

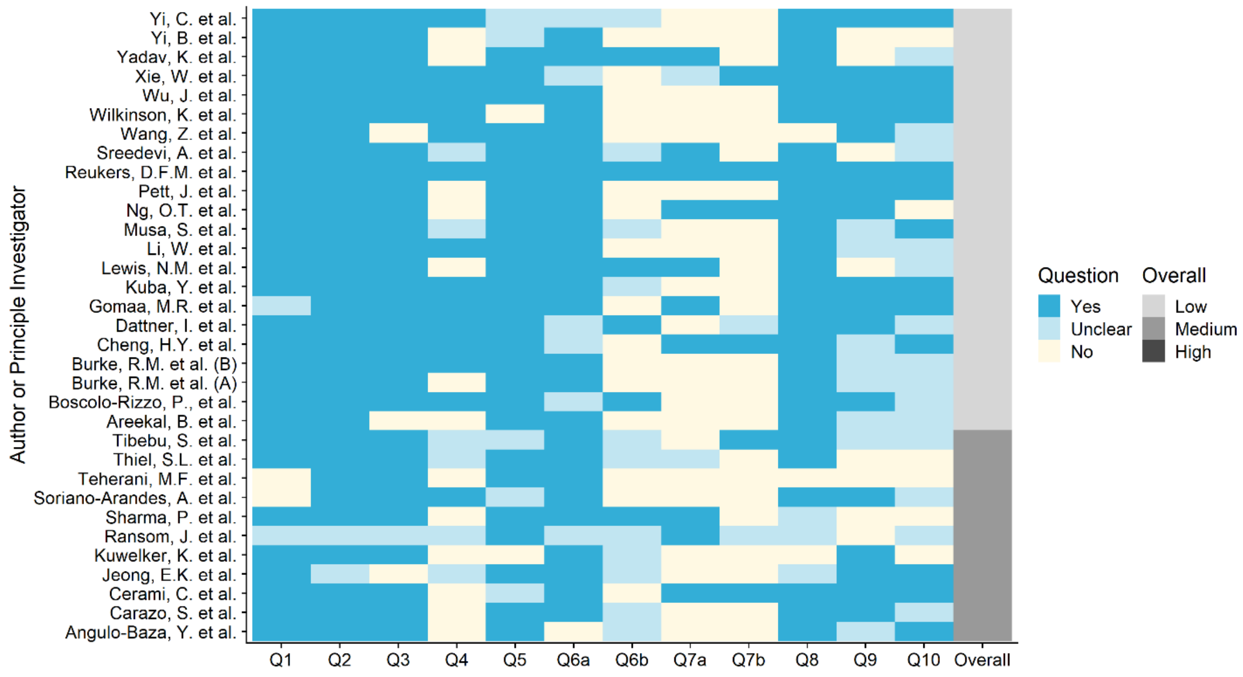


**Supplementary Figure 3:** Results of the critical appraisal tool as applied to investigations that reported household secondary clinical attack rate (hSCAR). Colours for Questions 1–10 indicate whether each was addressed in the investigation (dark blue) or not (cream), or instances where there was insufficient detail available to assess (light blue). An overall rating of the risk of bias is provided in the far-right column, with investigations rated Low (light grey), Medium (medium grey) or High (dark grey).

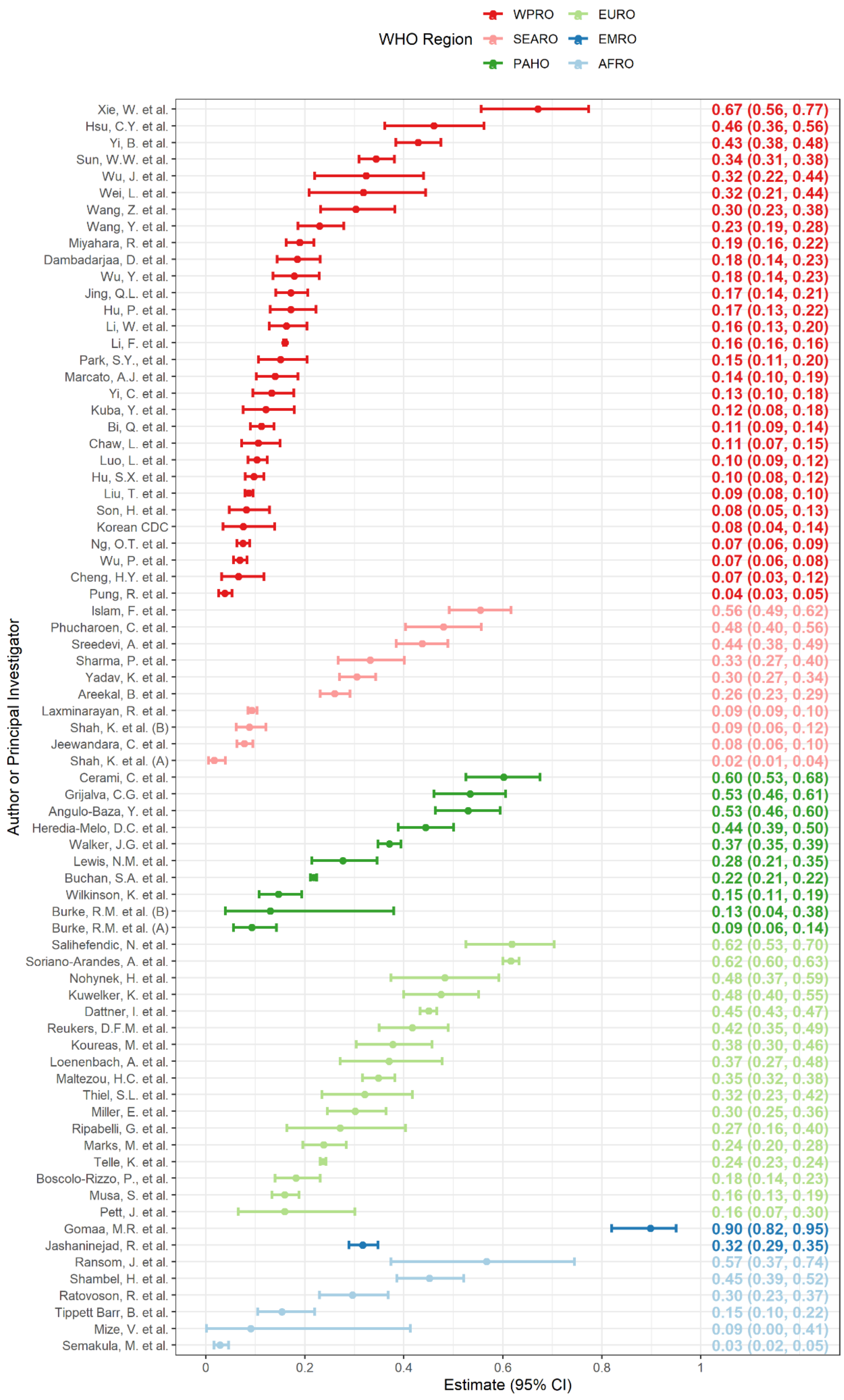


**Supplementary Figure 4.** Forest plot of estimated household secondary infection attack rates (hSAR) coloured by WHO Region. The estimated hSAR and 95% confidence interval are shown on the right margin.

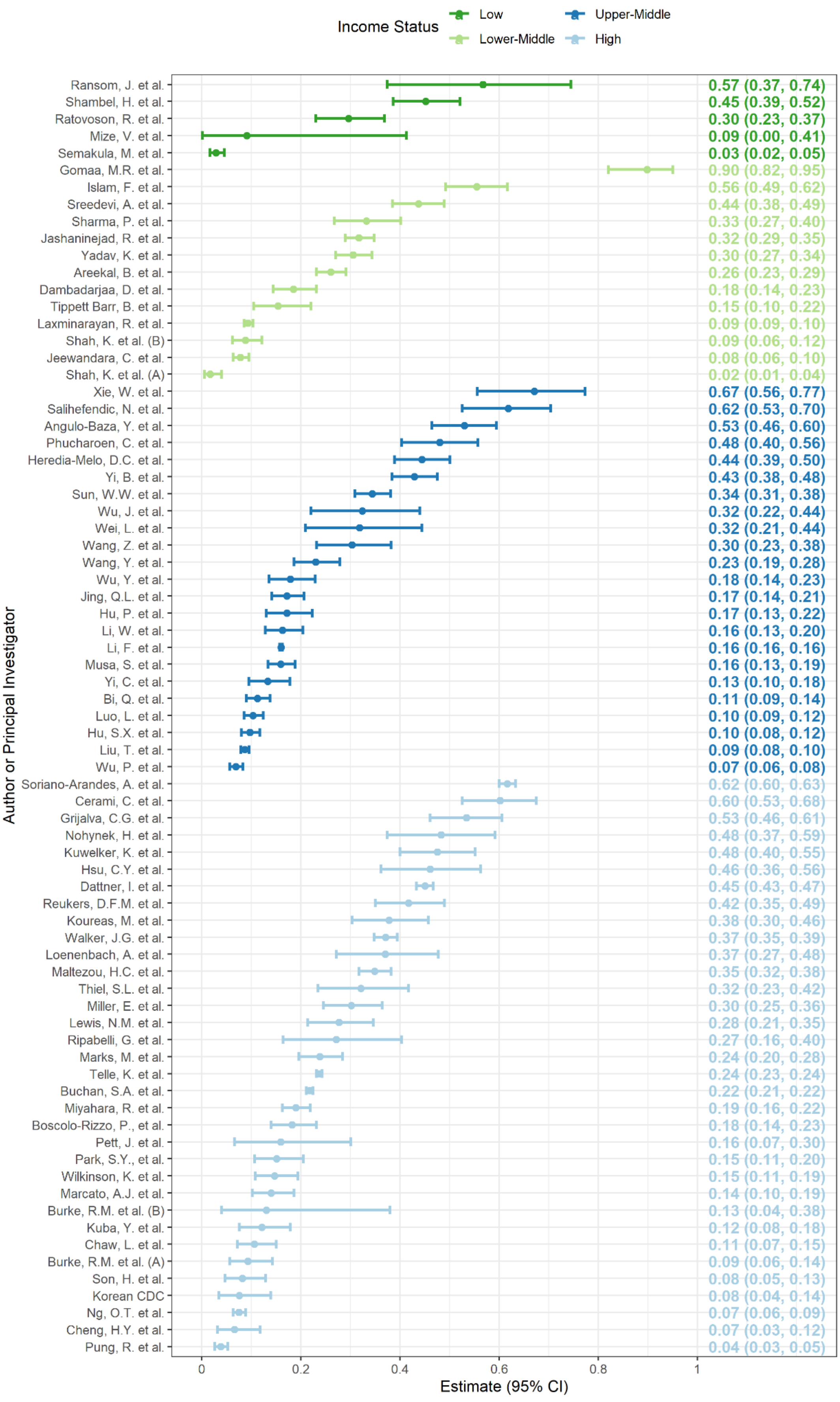


**Supplementary Figure 5.** Forest plot of estimated household secondary infection attack rates (hSAR) coloured by income status as reported by the World Bank in 2021.[81] The estimated hSAR and 95% confidence interval are shown on the right margin.

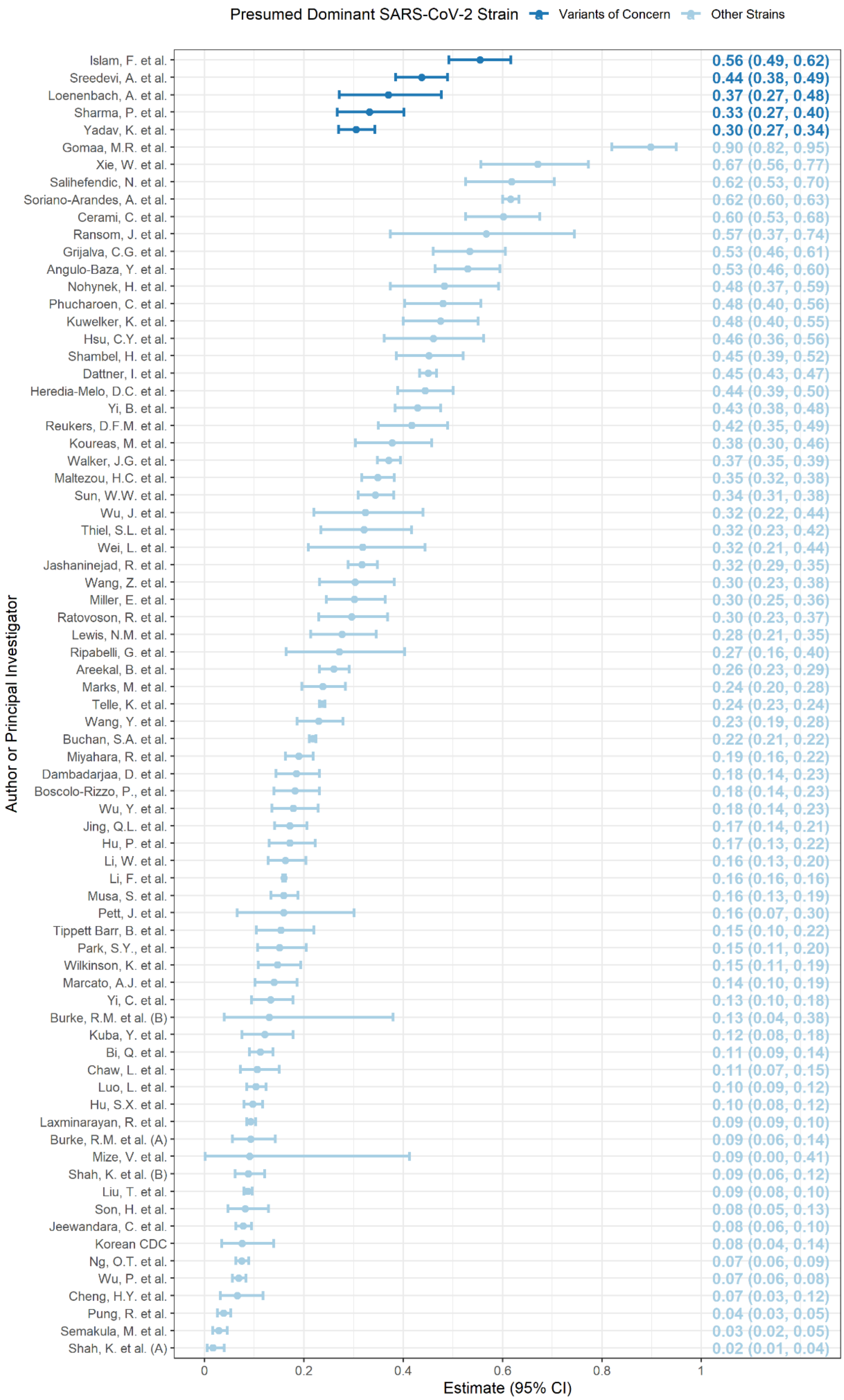


**Supplementary Figure 6.** Forest plot of estimated household secondary infection attack rates (hSAR) coloured by presumed dominant SARS-CoV-2 strain as determined by data available from CoVariants[82] GISAID.[83] The estimated hSAR and 95% confidence interval are shown on the right margin.

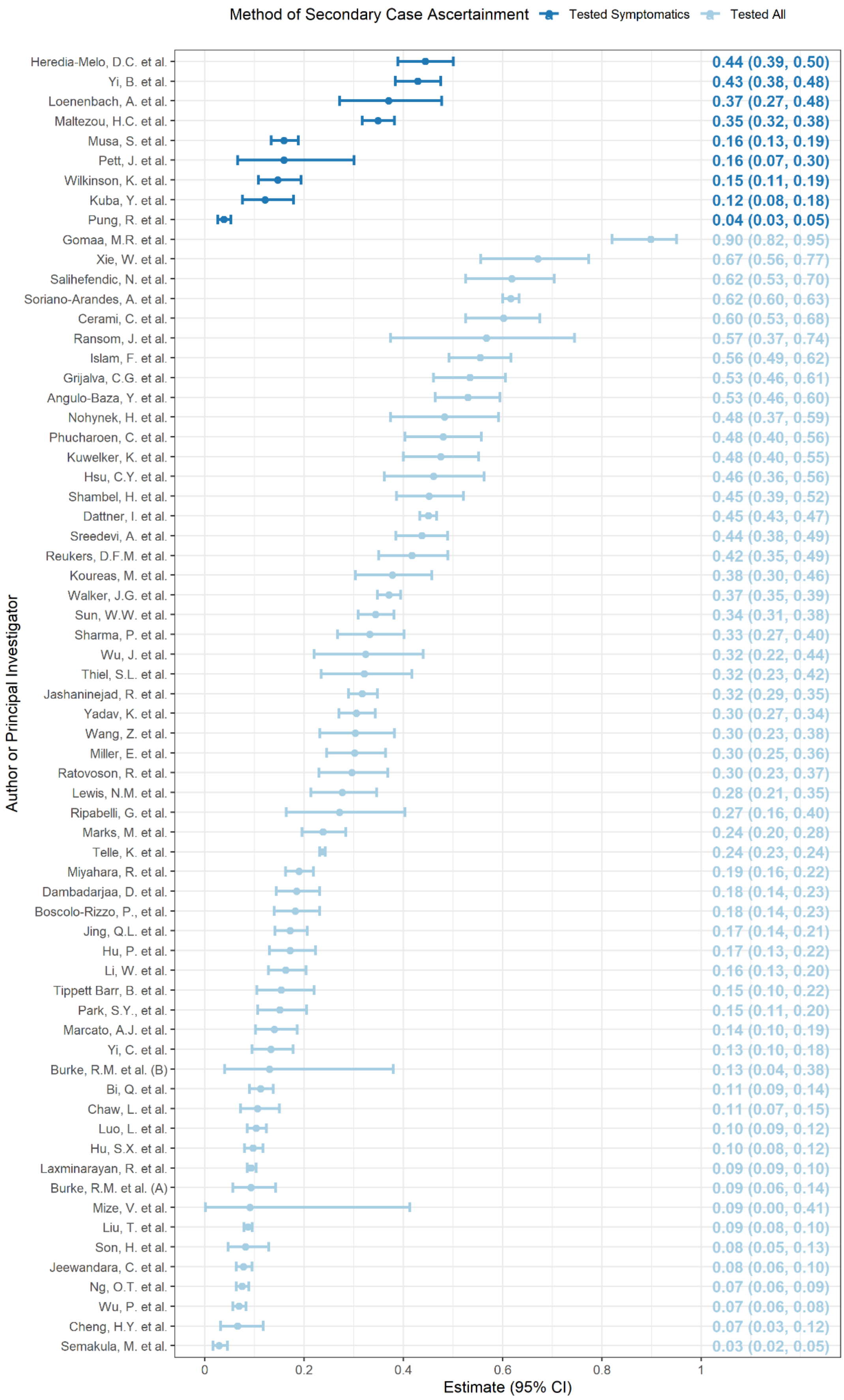


**Supplementary Figure 7.** Forest plot of estimated household secondary infection attack rates (hSAR) coloured by household contact testing protocol implementation. The estimated hSAR and 95% confidence interval are shown on the right margin.

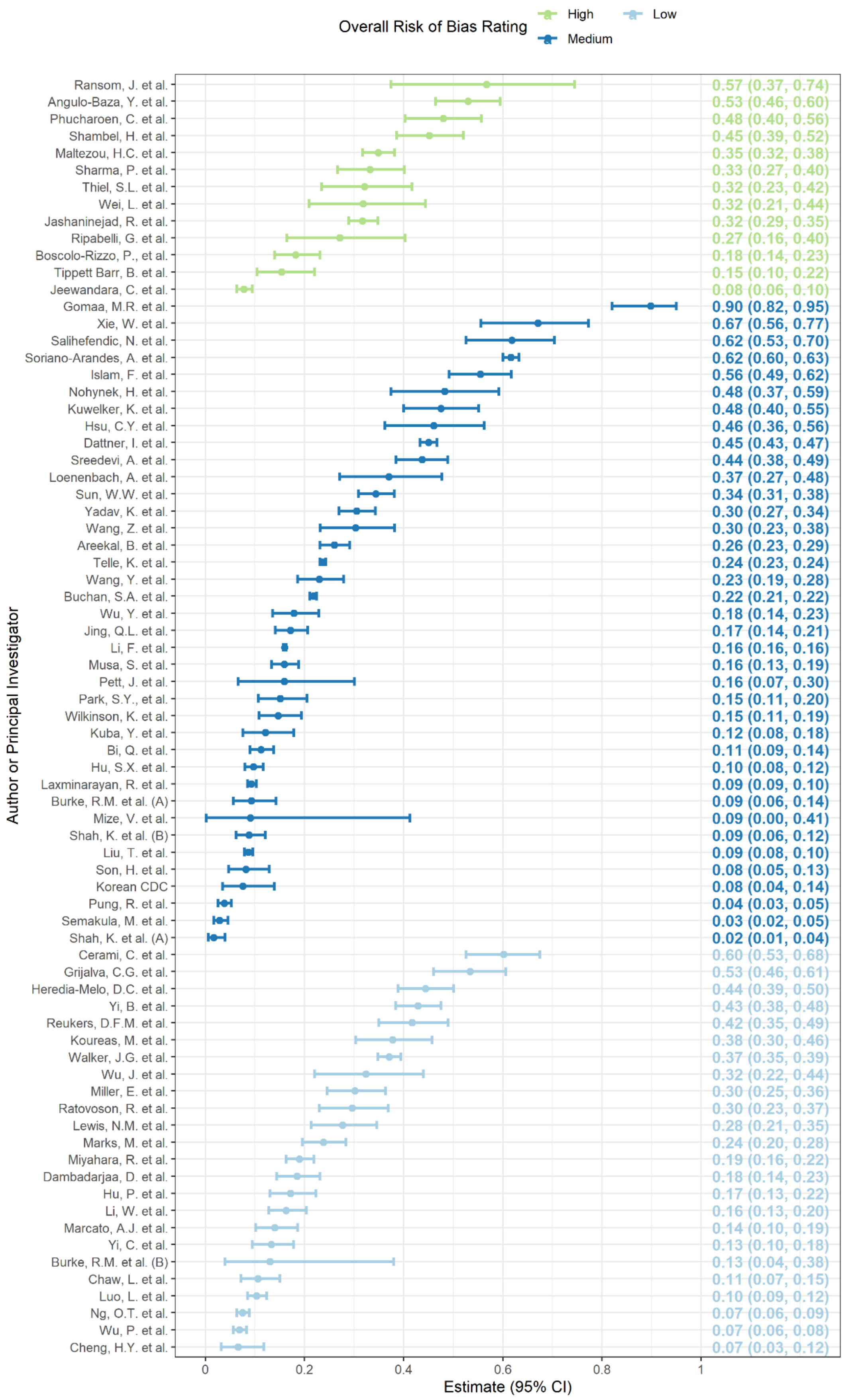


**Supplementary Figure 8.** Forest plot of estimated household secondary infection attack rates (hSAR) coloured by overall risk of bias assessment. The estimated hSAR and 95% confidence interval are shown on the right margin.

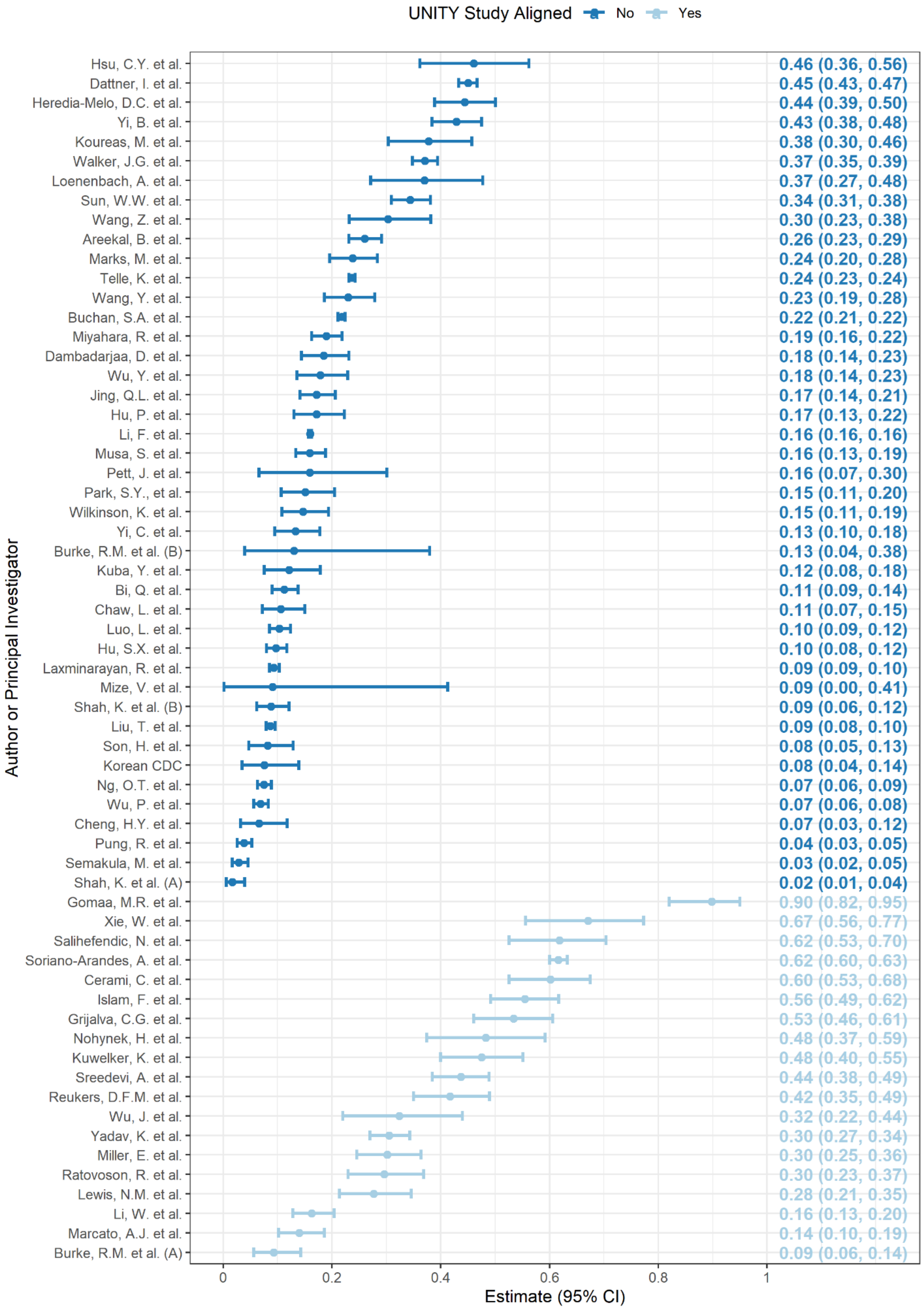


**Supplementary Figure 9.** Forest plot of estimated household secondary infection attack rates (hSAR) coloured by alignment to the UNITY protocol, as determined by household transmission study design, prospective data collection and routine testing of all household contacts. The estimated hSAR and 95% confidence interval are shown on the right margin.

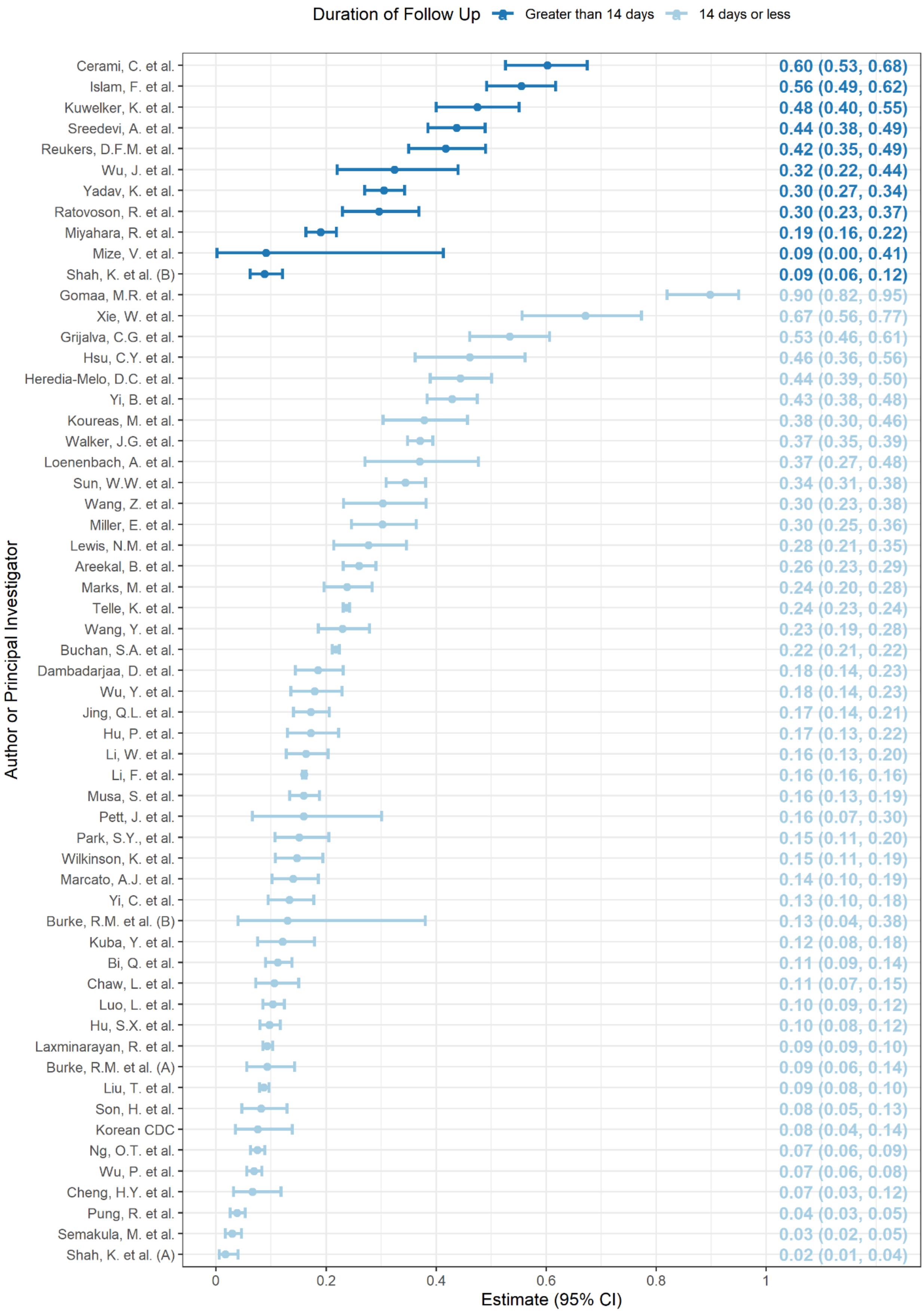


**Supplementary Figure 10.** Forest plot of estimated household secondary infection attack rates (hSAR) coloured by duration of follow up of household contacts. The estimated hSAR and 95% confidence interval are shown on the right margin.

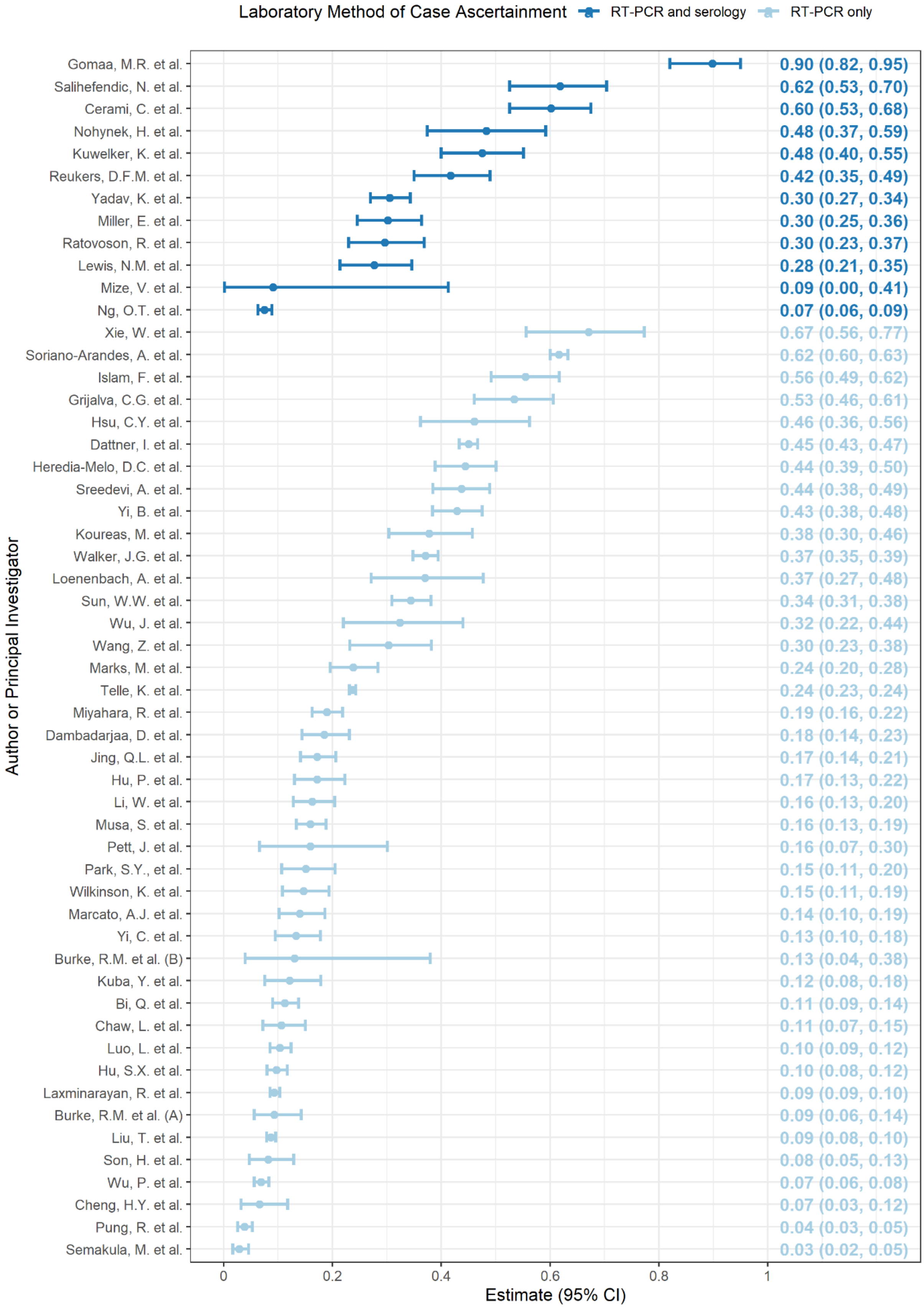


**Supplementary Figure 11.** Forest plot of estimated household secondary infection attack rates (hSAR) coloured by laboratory method of secondary case ascertainment. The estimated hSAR and 95% confidence interval are shown on the right margin.

**Supplementary Table 2.** Results comparing primary meta-analysis results to those from a risk of bias subgroup meta-analysis of household secondary infection attack rate (hSAR) after inclusion of studies at high risk of bias. $I^2$ and $\tau^2$ are presented for each model to indicate the percentage of variation across studies attributable to heterogeneity and the estimated between-study variance, respectively. The p-value from the $\chi^2$ test for heterogeneity is also presented.

| | No. studies | $I^2$ | $\widehat{\tau^2}$ | P-value |
|---|---|---|---|---|
| Infection household secondary attack rate | 62 | 99.7 | 1.190 | <0.0001 |
| **Pre-Specified Subgroup Analyses** | | | | |
| Risk of bias assessment | 75 | 99.6 | 1.069 | <0.0001 |
| *Low or moderate risk of bias* | 62 | | | |
| *High risk of bias* | 13 | | | |

**Supplementary Table 3**. Results from meta-analyses of household secondary clinical attack rate (hSCAR). $I^2$ and $\tau^2$ are presented for each model to indicate the percentage of variation across studies attributable to heterogeneity and the estimated between-study variance, respectively. The p-value from the $\chi^2$ test for heterogeneity is also presented.

| | No. studies | $I^2$ | $\widehat{\tau^2}$ | P-value |
|---|---|---|---|---|
| Clinical household secondary attack rate | 33 | 99.7 | 1.344 | <0.0001 |
| **Pre-Specified Subgroup Analyses** | | | | |
| Income Status | 33 | 99.7 | 1.324 | <0.0001 |
| *High income* | 19 | | | |
| *Low- and middle- income* | 14 | | | |
| Predominant circulating strain | 33 | 99.7 | 1.249 | <0.0001 |
| *Other strain* | 30 | | | |
| *Variant of concern* | 3 | | | |

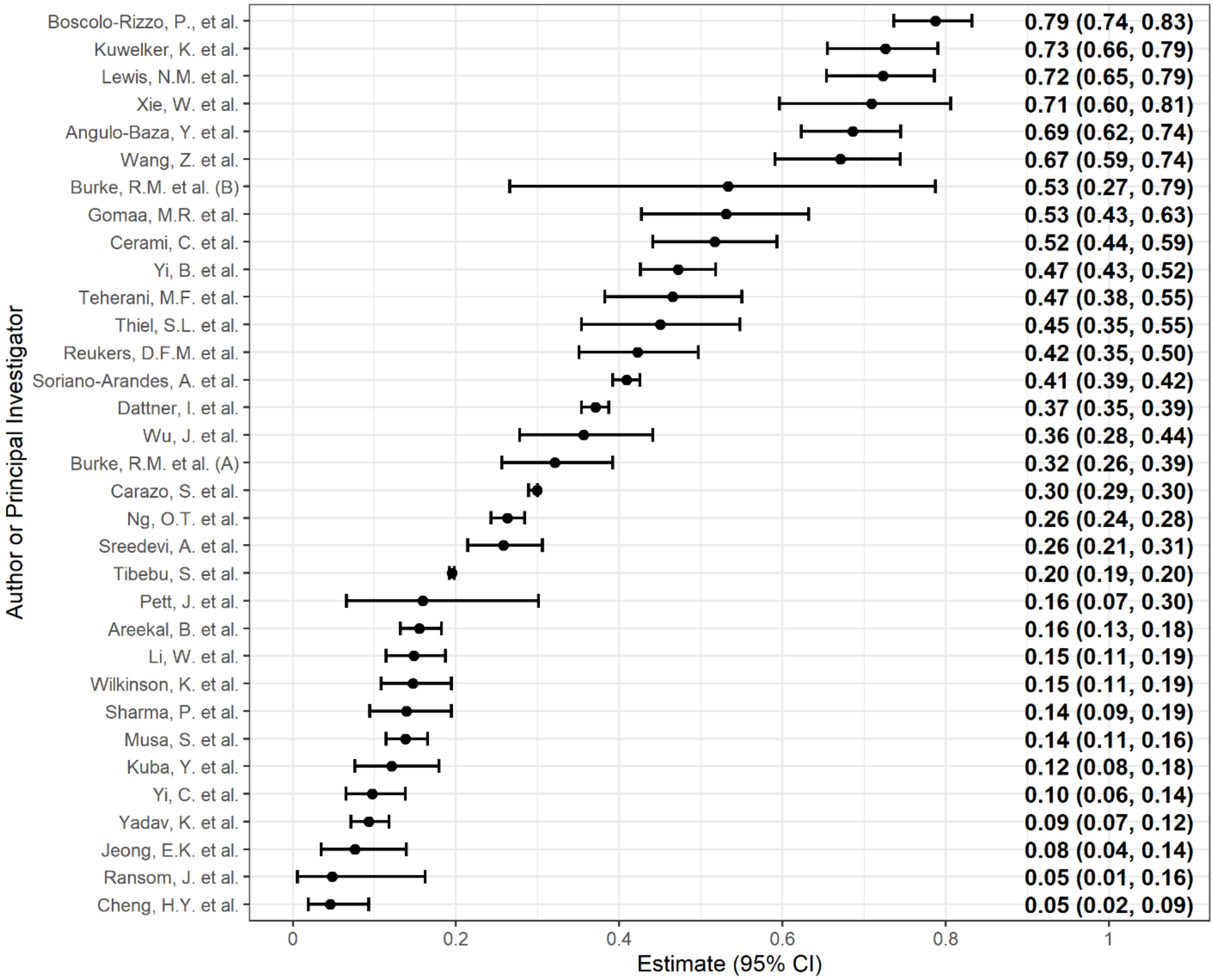


**Supplementary Figure 12**. Forest plot of the household secondary clinical attack rates (hSCAR) in included articles (n = 33), ordered from highest estimated hSCAR (top) to lowest estimated hSCAR (bottom). The hSAR and 95% confidence intervals (CI) are shown on the right margin.

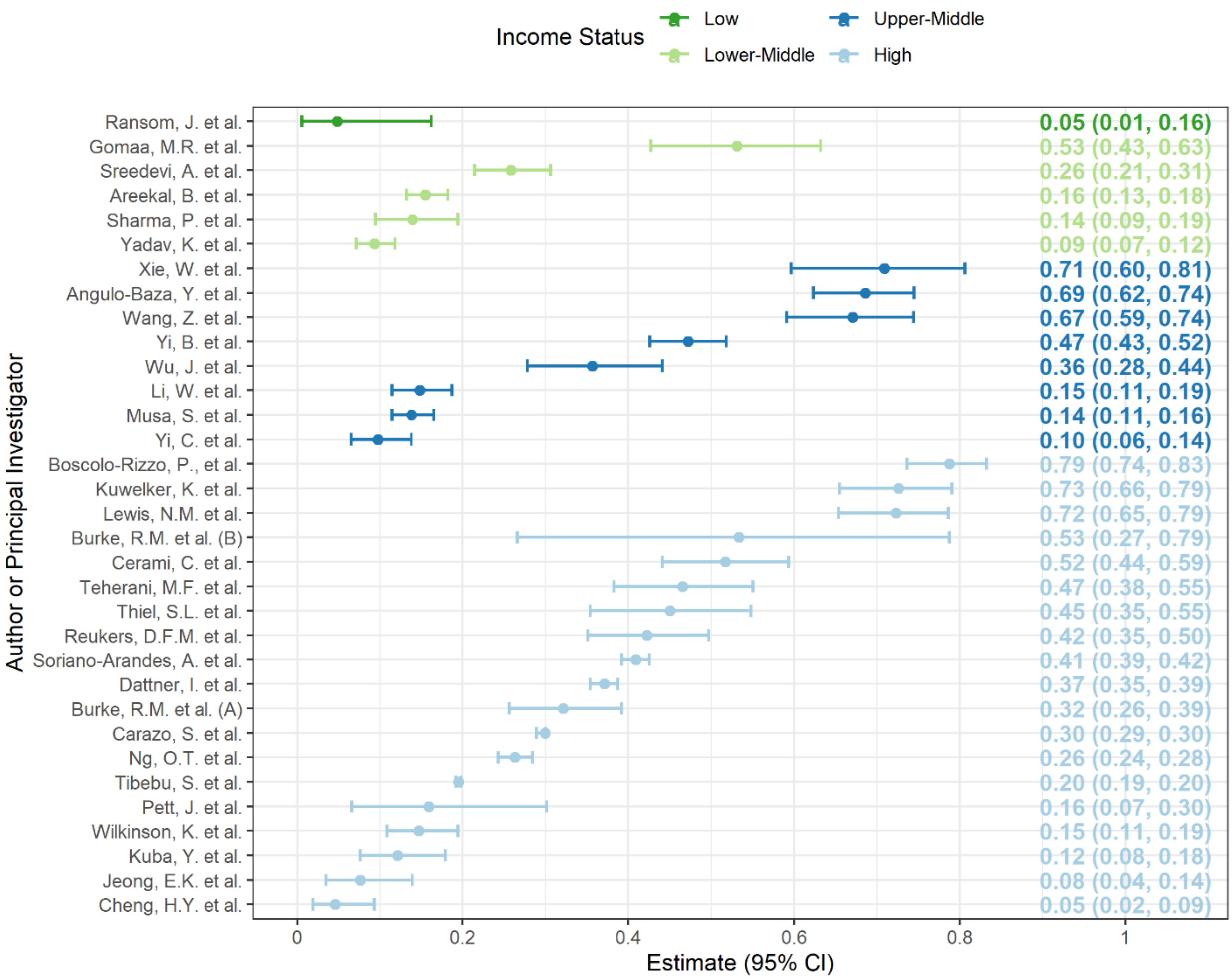


**Supplementary Figure 13.** Forest plot of estimated household secondary clinical attack rates (hSCAR) coloured by income status as reported by the World Bank in 2021.[81] The estimated hSCAR and 95% confidence interval are shown on the right margin.

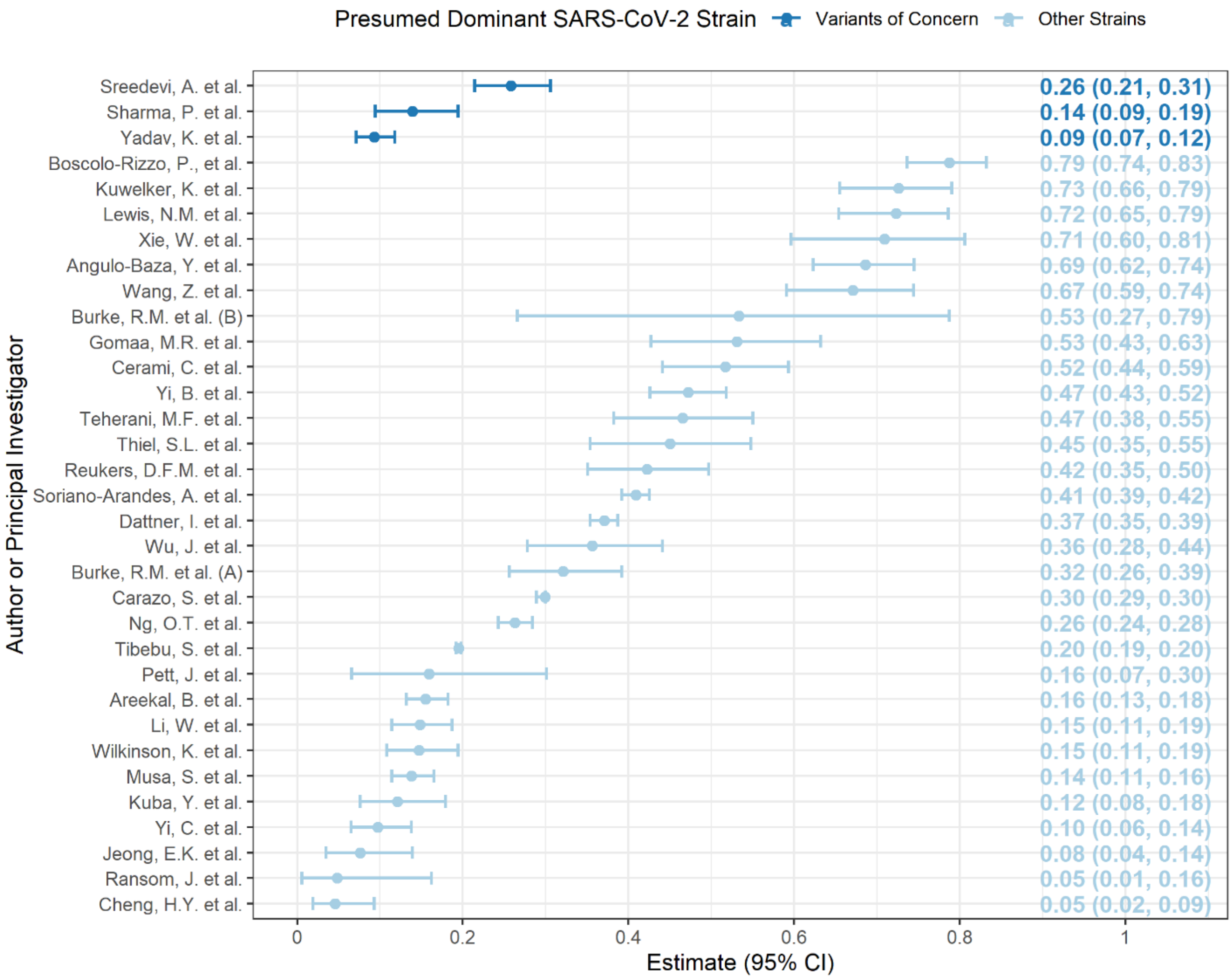


**Supplementary Figure 14.** Forest plot of estimated household secondary clinical attack rates (hSCAR) coloured by presumed dominant SARS-CoV-2 strain as determined by data available from CoVariants[82] and GISAID[83]. The estimated hSCAR and 95% confidence interval are shown on the right margin.
